# Maternal influenza-like illness and neonatal health during the 1918 influenza pandemic in a Swiss city

**DOI:** 10.1101/2024.12.13.24318720

**Authors:** Mathilde Le Vu, Katarina L Matthes, Eric Schneider, Aline Moerlen, Irene Hösli, David Baud, Kaspar Staub

## Abstract

**Background:** Exposure to the 1918 influenza pandemic may have been associated with preterm birth (<37 weeks). Other outcomes, such as infant size or weight, have rarely been explored.

**Objective:** To estimate whether *in utero* exposure to maternal influenza-like illness (ILI) during the 1918 pandemic was associated with pregnancy outcomes, and whether associations varied depending on ILI timing or on fetal sex.

**Design:** Cross-sectional study using historical birth records.

**Setting:** Lausanne maternity hospital.

**Participants:** 2,177 singletons born during the pandemic.

**Measurements:** The impact of ILI on gestational age, stillbirth, and anthropometric measurements, adjusted on covariates in generalized linear models. Analyses were stratified by fetal sex.

**Results:** 282 women developed ILI during pregnancy. ILI exposure was associated with lower anthropometric measurements: the odds ratio (OR) of low birth weight (<2,500g) was 2.06 [95%CI 1.33; 3.20]. In multivariable models, there was strong evidence that third trimester exposure was associated with adverse pregnancy outcomes, including with a higher preterm birth rate (OR 2.87 [95%CI 1.53; 5.39]). There was moderate evidence that first-trimester ILI exposure was associated with lower anthropometric measurements, in univariable models only. The magnitude of the declines in anthropometric parameters was higher among male fetuses, and they had a higher stillbirth risk. Only 41% of infants exposed to first-trimester ILI were males.

**Limitation:** our findings may not generalize to the entire population of Lausanne, as 34% of births were homebirths at the time.

**Conclusion:** maternal ILI may have triggered premature birth. The low sex ratio at birth for first-trimester exposure may indicate selection against males through miscarriage, but males were still more vulnerable than females to third trimester exposure.

**Primary funding source:** Swiss National Science Foundation.

## Introduction

The 1918 influenza pandemic had the unique feature of a high mortality rate among young adults (20-40 years old) ^1^. Pregnant individuals had a particularly high risk of developing severe symptoms, of being admitted to the hospital and of dying when contracting influenza ^2–6^; this was also the case during the recent influenza pandemic of 2009, albeit to a lower extent ^7–9^.

While maternal illness during the pandemic of 2009 was associated with higher rates of stillbirth and low birth weight (LBW, birth weight <2500g) ^10^, there are limited data on pregnancy outcomes during the 1918 pandemic. A contemporary study from 1919 noted that as many as 50% of 1,350 pregnant women with influenza-like illness (ILI) experienced pregnancy complications, including early pregnancy loss and preterm birth ^2^. Such events were more frequent if the mother had developed pneumonia ^3,5,6,11^. Population rates of stillbirth and neonatal mortality spiked during the pandemic in the UK and Japan ^12,13^, though it is uncertain whether it was due to *in utero* exposure to the virus, to maternal stress in times of crisis, or to the repercussions of the First World War. A short-term decrease in birth weight and placenta weight were reported in Basel, Switzerland in 1918-1919, coinciding with *in utero* exposure to both the pandemic and a food supply crisis towards the end of the First World War ^14^. Other neonatal anthropometric parameters have rarely been documented during the 1918 pandemic.

We take advantage of high-quality archival birth records from the city of Lausanne, in the Western canton of Vaud, Switzerland. The first pandemic wave reached the country in July 1918, followed by the deadliest wave from October 1918 to March 1919, and a milder wave in early 1920. We aim to estimate whether *in utero* exposure to maternal ILI was associated with higher risks of adverse pregnancy outcomes, reporting on infant size (including birth weight and head circumference), as well as preterm birth and stillbirth rates. To our knowledge, this is the first study - apart from contemporary physician reports - examining the direct effect of maternal ILI during the 1918 pandemic on pregnancy outcomes, while controlling for important covariates. Since the literature is inconclusive on the importance of the trimester during which ILI occurs ^15,16^, and because symptoms severity might be playing a role ^3,5,6,11,17^, we account for both the timing and the severity of ILI in our analyses. Furthermore, analyses are stratified by fetal sex, as males may be more vulnerable to adverse *in utero* environment than female fetuses ^18–21^.

## Methods

The birth registers of the University Maternity Hospital of Lausanne from 1905-1924 ^22^ were transcribed from the Cantonal archives of Vaud. Although only data from the pandemic time period are used in the analyses, we describe maternal and infant characteristics during 1905-1924, for comparison. Only singletons and events clearly qualified as “deliveries” on the maternity form were transcribed. Events qualified as abortions are reported separately (“miscarriages” section).

The following variables were transcribed (and in some cases, categorized): date of birth, date of last menstrual period, maternal age (years), height (cm), address, gravidity (number of pregnancies regardless of the outcome, categorized as 1, 2, >2), civil status (married or single/missing), occupation, syphilis status, neonatal sex (male or female), living status at birth (stillbirth or livebirth), birth weight (grams, g), head circumference (cm), birth length (cm), placenta weight (g), gestational age (GA) assessed at birth. Based on living status at delivery discharge, early neonatal mortality in the five days following delivery was reported. If there was a discrepancy between GA assessed at birth and GA based on date of last menstruation, or if the later was missing (*n*=630), we used GA assessed at birth (this process is described in the Supplementary material of a previously published work ^23^). From this calculated GA variable, we defined preterm birth (<37 weeks). Based on maternal residential address, a variable “living inside Lausanne” (yes or no/unsure) was defined. From qualitative information in a “general health status” section, we created a categorical morphology variable (obese, thin or neither), and the binary variables goitre and rickets. Maternal occupation was coded using the Historical International Standard of Classification of Occupations (HISCO) database ^24,25^. HISCO codes were then grouped into 3 classes, with class 1 and 3 respectively representing the lowest and highest socio-economic classes. Seasonality was categorized based on season of birth.

### Miscarriages

Events qualified as “abortions” (thought to reflect what would today be considered as miscarriage/early fetal loss - mean GA was 13.7 weeks) between 1909 and 1921 (n=920) were transcribed. These years were selected so that enough years surrounding the pandemic were available for comparing miscarriages rates.

Infants *in utero* during the pandemic (based on its course in *Figure S1*, those born after 01/07/1918 and conceived before 01/04/1919, or born after 01/01/1920 and conceived before 01/04/1920), and whose mother had ILI during pregnancy, were considered as exposed to ILI (n=282). As the record of illness is only based on symptoms the term ILI is used rather than “infected”. Disease onset was categorised by trimester: first (months 1-3), second (months 4-6), and third (from month 7 to delivery). Trimester of illness was available for 268 (95%) of the ILI cases. Symptom severity was classified as severe (bronchopneumonia, pneumonia, or bronchitis) or mild and was available for 237 (84%) of the ILI cases.

Investigated outcomes were birth weight, head circumference, placenta weight, birth length, ponderal index 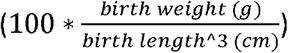, GA, LBW, preterm birth, stillbirth and early neonatal mortality in the first five days after delivery.

### Exclusion criteria

From the 13,042 births between 1905 and 1924, we successively excluded homebirths (*n*=52, 0.40%) due to missing information, infants with birth weight <500g or GA <22 weeks (*n*=13, 0.10%), based on modern definition for stillbirth ^26^, and outliers of birth weight, birth length, placenta weight and head circumference (*n* =11, 0.08%). The final dataset consists of 12,966 births. For the analyses, the dataset was restricted to the pandemic period, with infants born after 01/07/1918 and conceived before 01/04/1919, or born after 01/01/1920 and conceived before 01/04/1920 (n=2,177).

### Statistical analyses

Wilcoxon rank sum, Pearson’s Chi-squared and Fisher’ exact tests were performed for comparing exposed vs. unexposed groups. We fitted generalized linear models (GLMs), using a Gaussian family for the continuous outcome models and a logistic family with a log link for the binary outcome models. Univariable GLMs were built to model the relationship between maternal ILI and birth weight, head circumference, placenta weight, birth length, ponderal index, GA, LBW, preterm birth and early neonatal mortality among livebirths only (*n* =2,085). For the stillbirth outcome model, all births were used (*n* =2,177). Multivariable GLMs were built for the outcomes birth weight, head circumference, placenta weight, birth length, ponderal index, GA, LBW and preterm birth, and were adjusted for ILI, seasonality, maternal height, age, gravidity, neonatal sex, morphology, civil status, living inside Lausanne. All models were also fitted using a categorical variable describing the timing of maternal ILI, or using a categorical variable describing the severity of symptoms. The cases with unknown ILI timing or severity were excluded from GLMs. All models were stratified by fetal sex. However, for the models with ILI timing or severity, sex-stratified models were univariable only, due to the small number of events.

Regarding miscarriages, only descriptive statistics were used to compare maternal characteristics and yearly changes.

Additional information about the pandemic in Lausanne and Switzerland, and about methods are available in Supplementary material.

## Results

Deliveries at the maternity hospital rose from 315 in 1905 to 1,128 in 1924 (*Figure S2*), and it was associated with increases in maternal age, height, married women and women from the surroundings of Lausanne (Table S1, Figure S3). Overall, rates of LBW, preterm birth, stillbirth and early neonatal mortality declined through time, albeit with noticeable fluctuations (*Table 2, Figure S4*). During the pandemic, most mothers who delivered at the hospital were living outside of Lausanne, were married and multiparous (*Table S3*). As the majority of women were housewives, they belonged to the HISCO class 2, medium-skilled workers.

### Direct effect of maternal ILI

There were 282 (15%) mothers who developed ILI during the pandemic, and they had similar characteristics than those who did not (*Table S4*). The timing of ILI during pregnancy was consistent with the pandemic waves (*Figure S1*), with a peak of 47 women affected in October 1918 (*Figure S5*).

#### 1. Pregnancy outcomes depending on maternal ILI

Infants exposed to maternal ILI *in utero* had a lower birth weight, head circumference, placenta weight, birth length and ponderal index, and higher LBW and preterm birth rates (**Table 1**). More females (54%) were born to ILI-affected mothers. The magnitude of the impact of ILI on pregnancy outcomes was more important among males. For instance, there was stronger evidence of lower placenta weight among males than among females. In addition, there was strong evidence that head circumference was lower for males exposed to ILI, but no evidence for females.

**Table 1:** pregnancy outcomes depending on maternal ILI (during pregnancy and during the pandemic) and infant sex. *p: p-value*. ^1^Mean (SD); n (%). ^2^Wilcoxon rank sum test; Pearson’s Chi-squared test.

| ILI exposure | All births |  |  | Males |  |  | Females |  |  |
| --- | --- | --- | --- | --- | --- | --- | --- | --- | --- |
|  | no, N = 1,895<sup>1</sup> | yes, N = 282<sup>1</sup> | p-value<sup>2</sup> | no, N = 992<sup>1</sup> | yes, N = 130<sup>1</sup> | p-value<sup>2</sup> | no, N = 898<sup>1</sup> | yes, N = 152<sup>1</sup> | p-value<sup>2</sup> |
| birth weight (g) | 3,203.5 (562.4) | 3,083.0 (586.7) | <0.001 | 3,263.6 (584.8) | 3,099.7 (647.8) | 0.014 | 3,142.2 (520.0) | 3,068.8 (530.7) | 0.046 |
| head circumference (cm) | 34.4 (1.8) | 34.1 (1.9) | 0.003 | 34.7 (1.8) | 34.2 (2.1) | 0.006 | 34.1 (1.7) | 34.0 (1.8) | 0.4 |
| missing | 32 | 5 |  | 18 | 3 |  | 13 | 2 |  |
| placenta weight (g) | 586.6 (119.2) | 550.2 (107.4) | <0.001 | 595.7 (124.0) | 550.6 (112.4) | <0.001 | 576.9 (112.8) | 549.9 (103.3) | 0.002 |
| missing | 26 | 3 |  | 15 | 2 |  | 11 | 1 |  |
| birth length (cm) | 49.1 (2.7) | 48.7 (3.0) | 0.009 | 49.5 (2.7) | 48.8 (3.5) | 0.043 | 48.8 (2.5) | 48.6 (2.5) | 0.2 |
| missing | 5 | 1 |  | 3 | 0 |  | 2 | 1 |  |
| ponderal index | 2.7 (0.3) | 2.6 (0.3) | 0.034 | 2.7 (0.3) | 2.6 (0.3) | 0.031 | 2.7 (0.3) | 2.7 (0.3) | 0.3 |
| missing | 5 | 1 |  | 3 | 0 |  | 2 | 1 |  |
| gestational age (weeks) | 39.2 (2.1) | 39.0 (2.4) | 0.2 | 39.3 (2.1) | 38.9 (2.7) | 0.6 | 39.2 (2.0) | 39.1 (2.0) | 0.2 |
| sex |  |  | 0.045 |  |  |  |  |  |  |
| Male | 992 (52%) | 130 (46%) |  |  |  |  |  |  |  |
| Female | 898 (48%) | 152 (54%) |  |  |  |  |  |  |  |
| missing | 5 | 0 |  |  |  |  |  |  |  |
| low birth weight (<2'500g) | 157 (8%) | 38 (13%) | 0.004 | 85 (9%) | 21 (16%) | 0.005 | 70 (8%) | 17 (11%) | 0.2 |
| preterm birth (<37 weeks) | 153 (8%) | 33 (12%) | 0.042 | 78 (8%) | 18 (14%) | 0.022 | 73 (8%) | 15 (10%) | 0.5 |
| stillbirth | 80 (4%) | 12 (4%) | >0.9 | 45 (5%) | 8 (6%) | 0.4 | 33 (4%) | 4 (3%) | 0.5 |
| neonatal mortality d1-5 | 37 (2%) | 6 (2%) | 0.8 | 17 (2%) | 2 (2%) | >0.9 | 20 (2%) | 4 (3%) | 0.8 |
| missing | 0 | 1 |  |  |  |  | 0 | 1 |  |

In multivariable models, maternal ILI remained associated with lower birth weight, placenta weight, birth length and ponderal index, and higher LBW odds-ratio (OR), but the magnitude of the impact was generally smaller, than in univariable models (**Figure 1**, *Table S5*). On the other hand, there was weak evidence that ILI exposure was associated with head circumference and preterm birth rate, but the direction of the effects was consistent. In univariable models stratified by sex, ILI exposure among males was associated with larger decreases of anthropometric measures than among females (**Figure 1**, *Table S5*).

**Figure 1:**
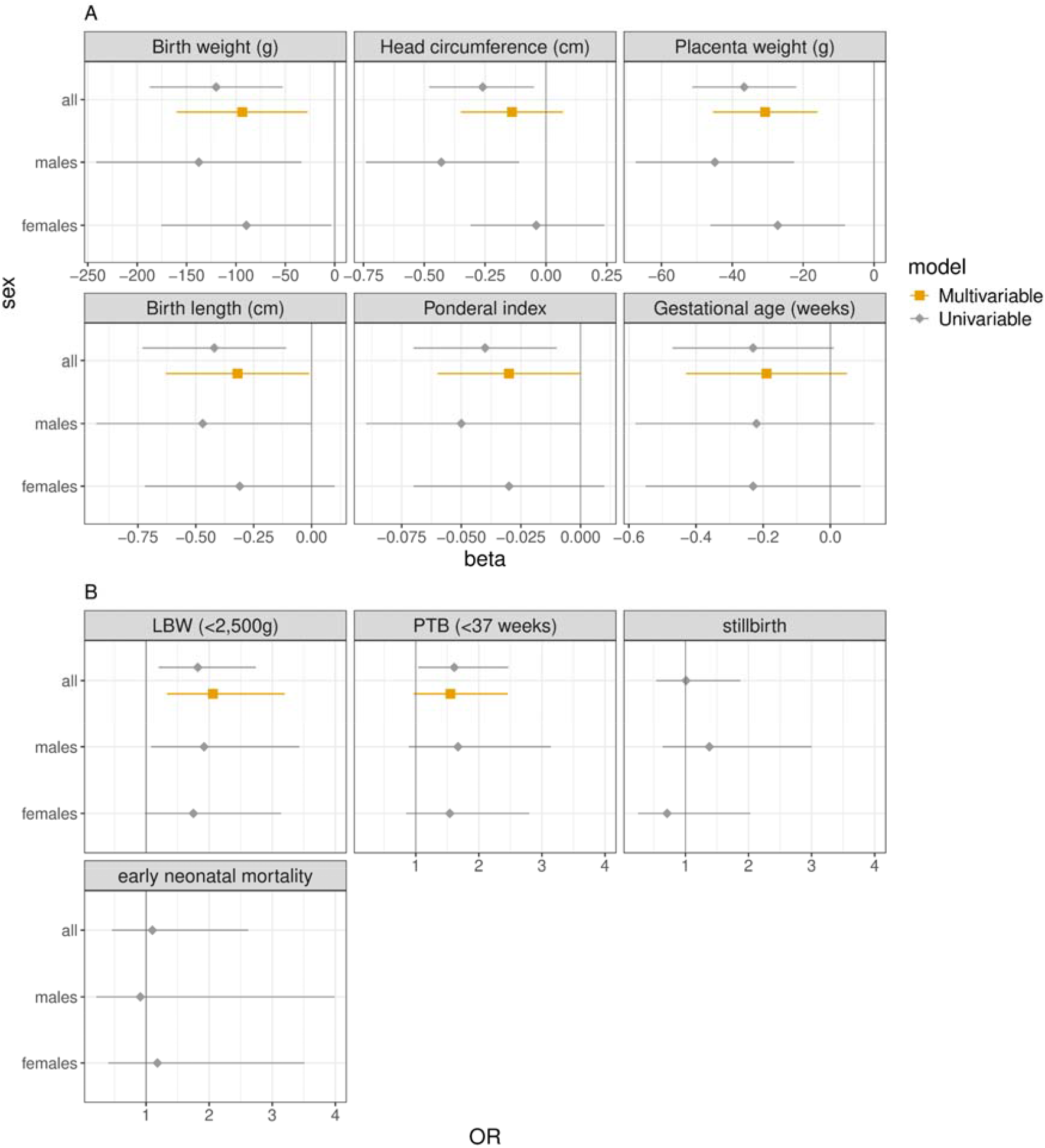
continuous (A) and binary (B) pregnancy outcomes depending on maternal ILI, univariable and multivariable GLMs, all births and stratified by sex. All models are fitted using only livebirths, except for stillbirth outcome. The same models are summarized in *Table S5*. Multivariable models are adjusted for: ILI during pregnancy, maternal age, height and gravidity, sex (except for the sex-stratified models), season, morphology, civil status, living inside Lausanne (details displayed in *Tables S6-8*).

#### 2. Trimester effect

Less women were affected by ILI in the first (*n*=75) than in the second (*n* =95) and third (*n* =98) trimesters. There were no major differences between maternal characteristics depending on the trimester of ILI (*Table S9*). Exposure in the third trimester was associated with lower infant anthropometrics measures than exposure earlier in pregnancy (**Table 2**). We note an effect of first trimester exposure on birth weight and birth length, but no evidence for second trimester exposure on pregnancy outcomes. There were more females born to ILI-affected mothers in the first (59%) than in the second (49%) and third (53%) trimesters. Interestingly, stillbirth risk was doubled when ILI occurred in the third trimester (OR 2.29, [1.12; 4.72]) (**Figure 2**, *Table S12*); this association could only be assessed in a univariable model, since there are few stillbirth events. In univariable GLMs (**Figure 2**, *Table S12*), exposure in the first and third trimesters were associated with lower infant size and weight, but the level of evidence was weaker for first compared to third-trimester exposure. In multivariable models, only third trimester exposure remained associated with poorer pregnancy outcomes (**Figure 2**, *Table S13*). For instance, the OR of LBW was 3.27 [95%CI 1.74; 6.16] and that of preterm birth was 2.87 [95%CI 1.53; 5.39]. Continuous GA was lowered for third trimester exposure to ILI (-0.79 weeks [95%CI -1.19; -0.39 weeks], *Table S13*).

**Table 2:**
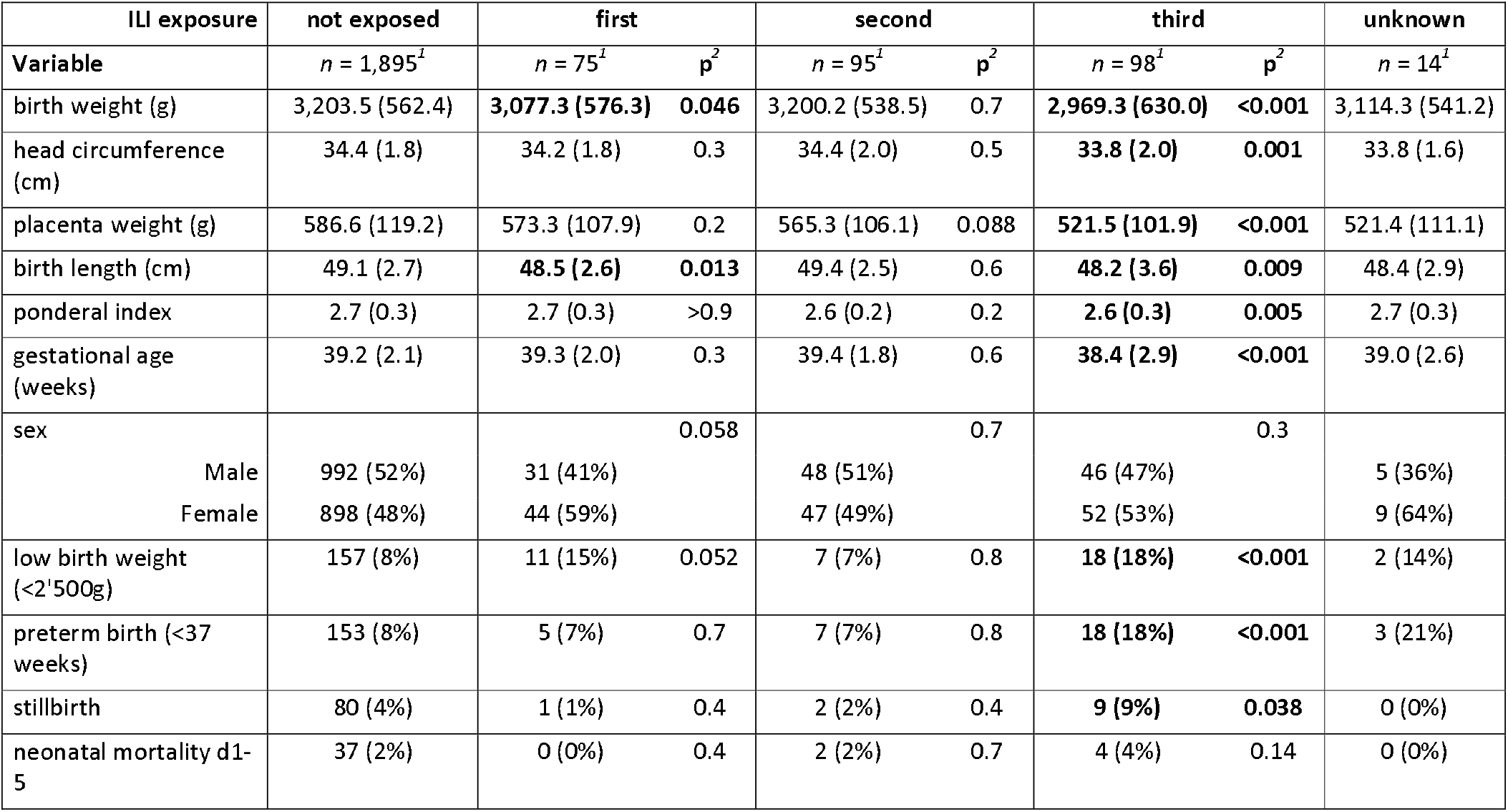
pregnancy outcomes depending on trimester of exposure to maternal ILI. The same tables separated for males and females are in *Tables S10-S11*. p: p-value. ^1^Mean (SD); n (%). ^2^Wilcoxon rank sum test; Fisher’s exact test. note: the p-value compares exposure to ILI during each trimester vs. not exposed.

**Figure 2:**
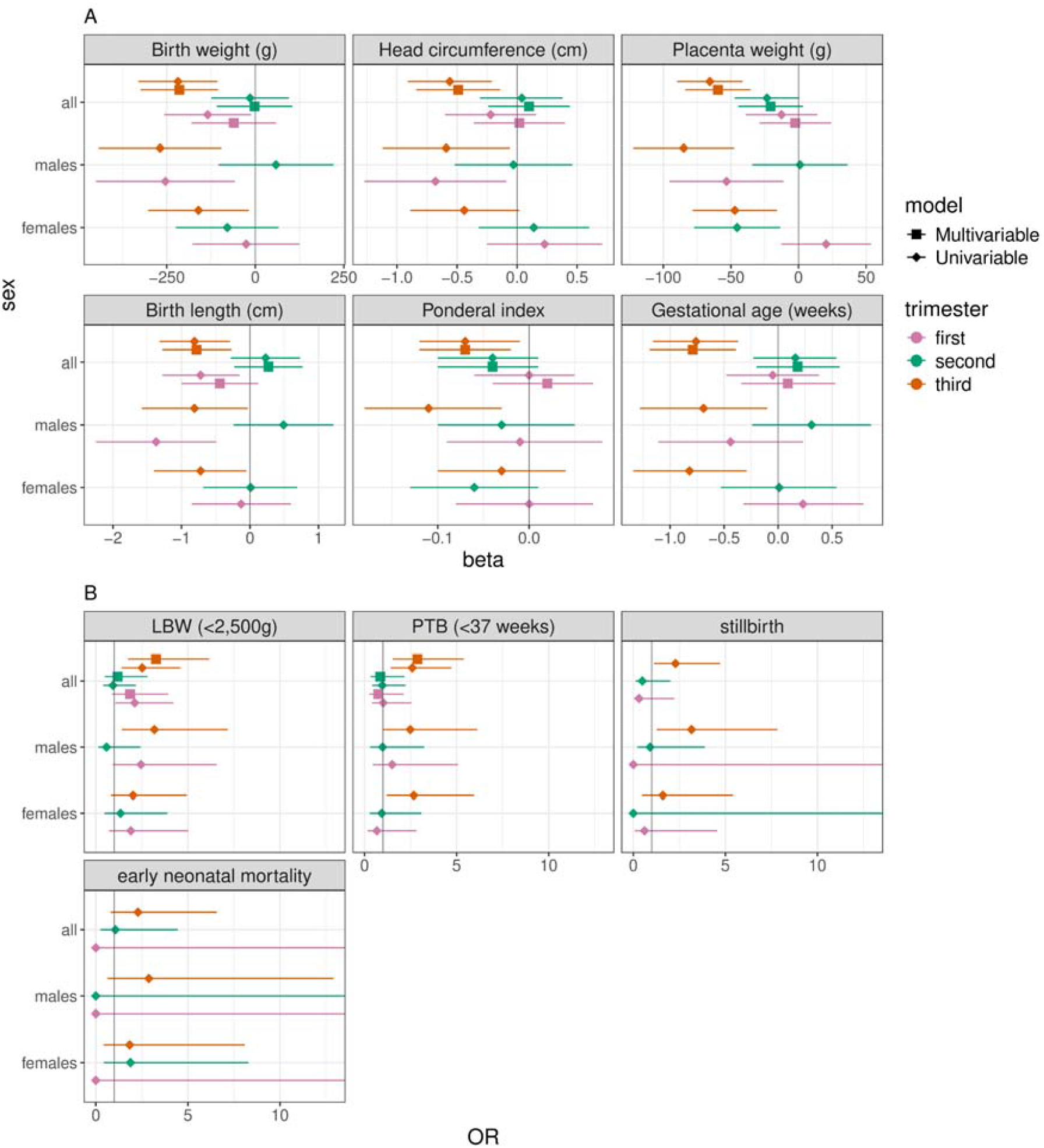
continuous (A) and binary (B) pregnancy outcomes depending on trimester of exposure to maternal ILI, univariable and multivariable GLMs, all births and stratified by sex. All models are fitted using only livebirths, except for stillbirth outcome. The same models are summarized in *Table S12-13*. For each outcome, only one GLM was fitted, and the exposure variable was a categorical variable (no ILI (reference category), ILI in the first, in the second, or in the third trimester). Note: when the timing of ILI exposure was unknown (*n*=14), cases were not considered.

In univariable models stratified by sex, only males were affected by first-trimester *in utero* exposure (**Figure 2**, *Table S12*). Females were affected by third trimester exposure, but generally to a lower extent that males were. For instance, birth weight was lowered by 269g [95%CI -443; -95g] for males and by 161g [95%CI -303; -18g] for females (**Figure 2**, *Table S12*).

#### 3. Severity of influenza-like illness symptoms

More women developed mild (*n*=137) than severe symptoms (*n*=100, **Table 3**). The only difference about mothers who developed severe symptoms was that more of them were multiparous (72%) compared to those who did not have ILI (60%) (*Table S14*). The more the pregnancy was advanced, the more often symptoms were severe: while only 15% of women who had ILI in the first trimester had severe symptoms, 49% of those affected in the third trimester reported severe symptoms (*Table S15*).

**Table 3:** pregnancy outcomes depending on the severity of ILI symptoms. The same tables separated for males and females are in *Tables S16-S17*. p: p-value. ^*1*^Mean (SD); n (%). ^*2*^Wilcoxon rank sum test; Fisher’s exact test. note: the p-value compares exposure to ILI with severe symptoms vs. not exposed.

| ILI exposure | not exposed | mild symptoms |  | severe symptoms |  | unknown symptoms |
| --- | --- | --- | --- | --- | --- | --- |
|  | *n* = 1,895<sup>1</sup> | *n* = 137<sup>1</sup> | *p*<sup>2</sup> | *n* = 100<sup>1</sup> | *p*<sup>2</sup> | *n* = 45<sup>1</sup> |
| birth weight (g) | 3,203.5 (562.4) | **3,092.4 (527.5)** | **0.008** | 3,089.0 (648.2) | 0.13 | 3,041.1 (624.8) |
| head circumference (cm) | 34.4 (1.8) | **34.1 (1.7)** | **0.021** | 34.2 (2.3) | 0.3 | 33.9 (1.9) |
| placenta weight (g) | 586.6 (119.2) | **550.7 (111.9)** | **<0.001** | **552.5 (102.9)** | **0.010** | 543.8 (105.2) |
| birth length (cm) | 49.1 (2.7) | 48.8 (2.7) | 0.068 | 48.8 (3.3) | 0.4 | 48.2 (3.0) |
| ponderal index | 2.7 (0.3) | 2.6 (0.3) | 0.10 | **2.6 (0.3)** | **0.063** | 2.7 (0.3) |
| gestational age (weeks) | 39.2 (2.1) | 39.2 (2.0) | >0.9 | 38.8 (2.7) | 0.12 | 38.9 (2.6) |
| sex |  |  | **0.033** |  | >0.9 |  |
| Male | 992 (52%) | **59 (43%)** |  | 53 (53%) |  | 18 (40%) |
| Female | 898 (48%) | **78 (57%)** |  | 47 (47%) |  | 27 (60%) |
| low birth weight (<2'500g) | 157 (8%) | 13 (9%) | 0.6 | **17 (17%)** | **0.003** | 8 (18%) |
| preterm birth (<37 weeks) | 153 (8%) | 15 (11%) | 0.2 | **14 (14%)** | **0.037** | 4 (9%) |
| stillbirth | 80 (4%) | 5 (4%) | 0.7 | 7 (7%) | 0.2 | 0 (0%) |
| neonatal mortality d1-5 | 37 (2%) | 1 (1%) | 0.5 | 4 (4%) | 0.14 | 1 (2%) |

In multivariable models, only placenta weight, ponderal index and LBW were still impacted by maternal ILI, with the latter two outcomes affected only when the mother developed severe symptoms (**Figure 3**, *Table S19*). In univariable GLMs stratified by sex, stillbirth rate was only higher among males exposed to a severe ILI (OR 2.69, [95%CI 1.09; 6.61]) (**Figure 3**, *Table S18*).

**Figure 3:**
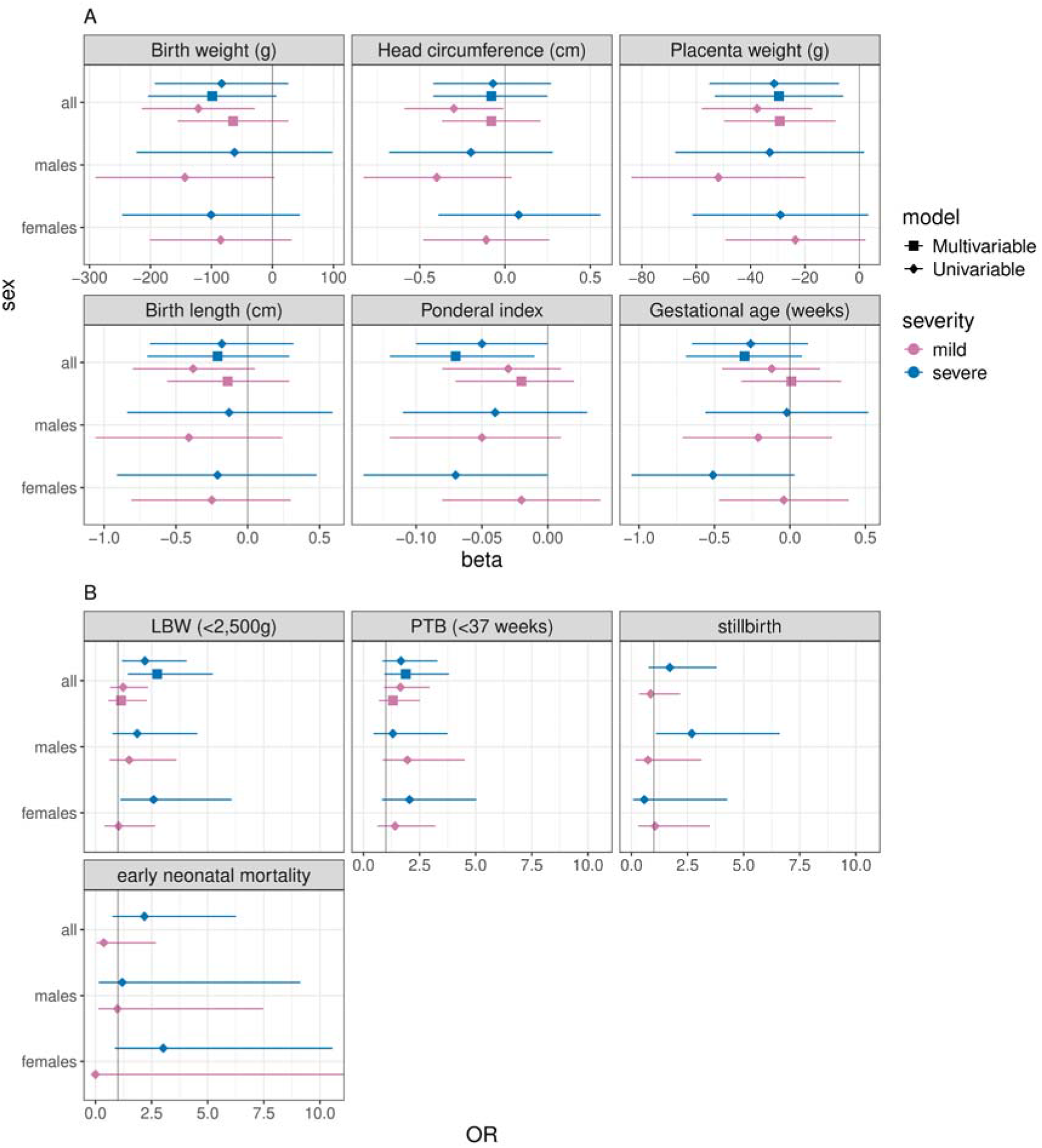
continuous (A) and binary (B) pregnancy outcomes depending on the severity of ILI symptoms, univariable and multivariable GLMs, all births and stratified by sex. All models are fitted using only livebirths, except for stillbirth outcome. The same models are summarized in *Table S18-19*. For each outcome, only one GLM was fitted, and the exposure variable was a categorical variable (no ILI (reference category), ILI with mild symptoms, ILI with severe symptoms). Note: when the severity of ILI was unknown (*n*=45), cases were not considered.

### Miscarriages

Yearly numbers of miscarriages peaked in 1918-1919 (*Figure S6A*), though percentages relative to births were also high before 1916 (*Figure S6E*). At the quarterly level, the miscarriage rate was already high before the pandemic (first quarter of 1918, *Figure S11F*). However, among women who miscarried in 1918, 1919 and 1920, 13%, 39% and 58% had ILI during pregnancy, respectively (*Table S20*). Comparing women who had miscarriages to those who delivered, we again see that ILI during pregnancy was more frequent among miscarriage cases (34% compared to 12%, *Table S21*).

## Discussion

### Infant health is negatively affected by maternal ILI, especially for third trimester exposure

Most measured anthropometric indicators were lowered by maternal ILI, and this was mainly driven by third trimester exposure. For instance, while birth weight was overall lowered by about 100g by maternal ILI, it was lowered by 215g when ILI occurred in the third trimester. In addition, infants whose mothers developed ILI in the third trimester were born on average 5.5 days earlier, with a preterm birth rate almost three times higher. First trimester exposure was associated with poorer pregnancy outcomes in univariable but not in multivariable models.

As GA is a strong determinant of fetal growth, being born earlier results in a lower birth weight and size. Thus, our findings of lower infant weight and size are probably - at least partly - mediated by shorter gestation. This is consistent with contemporary studies that reported early pregnancy loss and preterm birth among mothers with ILI ^2,3,5,6,11^. In the 1919 report carried out by the health authorities of Vaud, physicians frequently reported earlier deliveries, especially when illness occurred closer to the term ^27^. In a study using the same birth records than in the current paper, we reported lower birth weight and head circumference for *in utero* exposure to maternal ILI ^23^. However, GA was not affected: we believe this is because the timing of ILI was not considered. In addition, the dataset had not been restricted to the pandemic period but to the years 1911-1922, and as a result there were many more unexposed cases than in the current study (*n*=8,131 vs. *n*=1,895). During the 1957-1958 ^28^ and 2009-2010 ^17,29^ pandemics as well, there were reports of premature deliveries. In fact, even seasonal influenza may be associated with shorter gestation ^30,31^. In Denmark, seasonal flu was associated with lower birth weight only when GA was not controlled for ^31^. We chose not to adjust our models for gestational duration to avoid potential collider bias (*see Supplementary Methods*), but this Danish study supports our hypothesis that ILI-triggered premature delivery is the reason that birth weight is lower.

At Lausanne maternity, the proportion of women with ILI was higher among those who had a miscarriage compared to those who delivered. A high rate of miscarriage during the 1918 pandemic has long been assumed to be one of the reasons explaining the decrease in the number of births in the pandemic aftermaths ^32–37^. However, selection into fertility and especially deliberate postponement of conceptions during the pandemic may have also played a role, and we cannot conclude on the effect of ILI on miscarriage risk. Additionally, the miscarriage reports may not be representative of all events, as many can go unnoticed or occur at home without resulting in a hospitalisation.

### Timing of maternal ILI

There were less women with ILI in the first (27%) than in the second (34%) and third trimesters (35%). A systematic review concluded that during the 2009-2010 pandemic, more women were infected during the third trimester (47%) than in the first (9%) ^7^. Seasonal flu was also much more frequent in the third trimester ^31^. Infection closer to the term may lead to more severe symptoms, and would thus have a higher chance of being noticed. Indeed, we find that most severe cases occurred in the third trimester, with about 50% of women having developed pneumonia or bronchitis. Towards the end of pregnancy, altered immunity and reduced lung capacity increase the risk of severe influenza symptoms ^7,38^. Recall bias is another explanation, but first-trimester illness may also have been associated with miscarriages, and we selected women whose fetuses “survived” to first-trimester exposure.

### Males are more vulnerable to maternal ILI

The magnitude of the impact of maternal ILI was greater among males than females, with larger reductions in anthropometric weight and size. Furthermore, only males were affected by first-trimester exposure. Less males were born to mothers who experienced ILI in the first trimester compared to mothers who were not affected by influenza (41% vs. 52%). This may signal selection against males: if first trimester ILI led to miscarriage more frequently among males, then the sex ratio male:female at birth would be lower. Unfortunately, fetal sex was rarely reported among the miscarriage events and we cannot confirm this hypothesis. In Japan, the sex ratio at birth was also lower for first trimester exposure to the 1918 pandemic (indirectly measured using influenza deaths rates) ^21^. There is evidence that the sex ratio at birth decreases following other types of crisis, including natural disasters, economic crises, and terrorist attacks ^18,20,39,40^.

To conclude, it seems that males remained more vulnerable to influenza during the whole pregnancy, likely through a higher rate of miscarriage for first trimester exposure, while third trimester exposure was associated with lower size and weight measured at birth and a higher rate of stillbirth. Higher rates of miscarriage and stillbirth may have led to survival bias for surviving males at birth, but these effects appear to have been countered by the scarring effects of maternal ILI.

### Limitations

The definition of ILI is based on clinical symptoms reported by the mother, and they may be undistinguishable from that of other viruses, potentially leading to a misclassification of exposure. By focusing on illnesses occurring during the pandemic waves, we mitigate this bias and discard cases potentially due to seasonal flu or other diseases. Still, ILI diagnosis is limited to women seeking care or having more severe symptoms.

Finally, we do not know how representative the women who delivered at the maternity hospital are of all those who gave birth in Lausanne and its surroundings. Although there was only one maternity in the city, it was frequent to deliver at home ^41^. Based on official statistics, 66% of births occurring in Lausanne happened at the maternity in 1920 ^42^. In the same year, 10% of mothers delivering at the hospital were single, compared to 5% among homebirths. Paternal occupation indicated that socio-economic status was slightly lower among married mothers delivering at the maternity (data not shown). Still, there have never been restrictions on admission at the maternity ^43^, with both complicated and uncomplicated deliveries taking place, making us believe that our results are generalizable. Importantly, influenza seems to have affected all women, independently of socio-demographic characteristics, as reported in another paper ^44^.

### Implications

The poorer outcomes we report in the case of maternal ILI may have lasting consequences. Being born with a LBW is associated with a higher risk of morbidity in childhood ^45^, and with increased risks of chronic diseases in adulthood ^46–51^. Several health economics papers report that early-life exposure to the 1918 influenza pandemic negatively impacted socio-economic attainments and health outcomes later in life ^52–56^. This was also the case for the pandemic birth cohort in Switzerland, with lower educational attainment and occupational status ^57^. However, unobserved factors may be associated with both the poor *in utero* environment and long-term outcomes: we can hardly separate the cause of fetal stress from other factors which also generate poor health later in life. In addition, the negative long-term economic effects documented for the 1919 US birth cohort ^52^ were confounded by a negative socio-economic selection of parents ^58^.

In conclusion, our results highlight the adverse effects of maternal ILI during an influenza pandemic, based on over two thousand birth records. Our findings suggest that maternal ILI, particularly during the third trimester of pregnancy, may have triggered premature birth. The 1918 influenza pandemic was the most severe of the last century, and as influenza pandemics regularly occurred throughout history, they are bound to reoccur in the future. Our findings emphasize the importance of mitigating exposure to influenza during pregnancy through effective preventive measures.

## Data Availability

The data that support the findings of this study and the codes used for the data analysis will be made publicly available in the following online repository, after publication of the paper.

https://github.com/MathildeLV/Lausanne_influenza

## Acknowledgments

The authors would like to thank Veronika W Skrivankova, Frank Rühli, Sabine Rohrmann and Viktor von Wyl for their helpful comments, Urmila Bhattacharyya, Joel Floris, Antonia Galliker, Nora Haag, Rosa Hinselmann and Ellen Hünerwader for participating to the data transcription (along with authors Mathilde Le Vu, Katarina Matthes, Aline Moerlen and Kaspar Staub); the Cantonal archives of Vaud and its archivists for the access to the original data.

## Data Availability Statement

The data that support the findings of this study and the codes used for the data analysis will be made publicly available in the following online repository: https://github.com/MathildeLV/Lausanne_influenza, after publication of the paper.

