## Supplementary material for "Maternal influenza-like illness and neonatal health during the 1918 influenza pandemic in a Swiss city"

Historical context

*Socio-economic context*

From 1870, Swiss living standards markedly improved, interrupted only by the two World Wars (Floris et al., 2019). By the First World War, Switzerland was one of the three richest countries (among Western Europe and the USA), in terms of GDP per capita. Although Switzerland was not directly involved in the conflict, the country still suffered a long-lasting economic crisis (Floris et al., 2019). Nonetheless, in the majority of the period under study (1900s-1920s), Switzerland can be considered a high-income country. Infant mortality rates drastically declined between 1870 and the 1920s, at the national level and also in Lausanne, thanks to sanitation and modernisation of the water supply (Floris & Staub, 2019). The number of inhabitants in the Canton of Vaud was 317,457 in 1910 and 317,498 in 1920 (Jaeger, 1923). In the city of Lausanne, it was 64,446 in 1910 and 68,533 in 1920 (Jaeger, 1923).

*The maternity hospital in Lausanne*

In 1890, the Hospital of Lausanne became a university hospital, and it was the only one in the city. No restrictions were put into place at admission (Hôpital Cantonal de Lausanne, 1882). A new and larger building was built in 1916. As a result, an increasing proportion of births took place at the hospital.

*The 1918-20 influenza pandemic in Switzerland, the canton of Vaud and in the city of Lausanne*

The first pandemic wave hit the country in July 1918, while the second and deadliest one spread from October 1918 until March 1919. A third and milder wave took place between January and March 1920. More than half of the Swiss population was infected by influenza, and about 25,000 died of it in the first pandemic year alone (Sonderegger & Tscherrig, 2016).

In the early 20^th^ century, there were 25 federal member states in Switzerland, the so-called cantons. The city of Lausanne is located in the canton of Vaud; compared with the other Swiss cantons, the canton of Vaud was moderately affected by the pandemic and followed the wave pattern of Switzerland. It was estimated that 2,221 people died of the influenza, i.e. 0.70% of the population (Eidgenössisches Statistisches Bureau, 1919; Sonderegger, 1991). Half of them were men and 32% were aged 20-40 years old (Eidgenössisches Statistisches Bureau, 1919; Sonderegger, 1991). Compared to other Swiss cantons, the canton of Vaud introduced the mandatory reporting of influenza relatively late, on September 24^th^ 1918, i.e. after the summer wave (Koch, 2019; Messerli F-M, 1920). The Swiss Army was demobilized in November 1918 and a general strike followed, assumed to have caused the strong wave in the end of 1918 (Autorités de la santé du canton de Vaud, 1919). In Lausanne, during the first month of the pandemic (July 1918), there were already 101 deaths due to the flu. In comparison, the 1889 pandemic made only 2 victims in the first month (Autorités de la santé du canton de Vaud, 1919).

After first two waves of the pandemic in mid-1919, the health authorities of the canton of Vaud carried out a review of the pandemic (Autorités de la santé du canton de Vaud 1919). This was done within the framework of a detailed survey to all physicians in the canton. A hundred and eighteen physicians responded to the 15 questions about the course of the pandemic and the disease. These questionnaires were processed into a report. It was estimated that around 55% (*n* = 175,000) of the cantonal population had been infected (Autorités de la santé du canton de Vaud, 1919). There is a whole section on influenza among pregnant women: most doctors agreed that the flu was more dangerous for pregnant women, with higher frequencies of pulmonary complications. Early pregnancy loss and preterm birth was also noted. Dr Gilliard from Chateau d’Oex observed three cases in which both the mother and infant died. In Lausanne, Dr Zbinden observed frequent miscarriages and preterm births, while Prof Démiéville stated that flu among pregnant women was “extremely severe”. Dr Perrin, in Avenche reported 50% cases of miscarriages among pregnant women with influenza. In general, if the infection occurred close to the term of pregnancy, consequences were more severe for the mother and the child.

Incidence of influenza-like illnesses and hospitalisations in the canton of Vaud, as well as death counts in the city of Lausanne between 1916 and 1920 are displayed in *Figure S1*, using data from the Swiss public health weekly reports (Bühlmann, 1916).

**Figure S1: timing of the influenza pandemic in Lausanne and in the canton of Vaud.** Influenza-like illness incidence in the Canton of Vaud (**A**) and in the city of Lausanne (**B**); the dashed line indicates the introduction of the cantonal reporting obligation on 24.09.1918. All-cause deaths in the city of Lausanne (**C**); and hospitalisations in the canton of Vaud, due to infections (including influenza) (**D**).

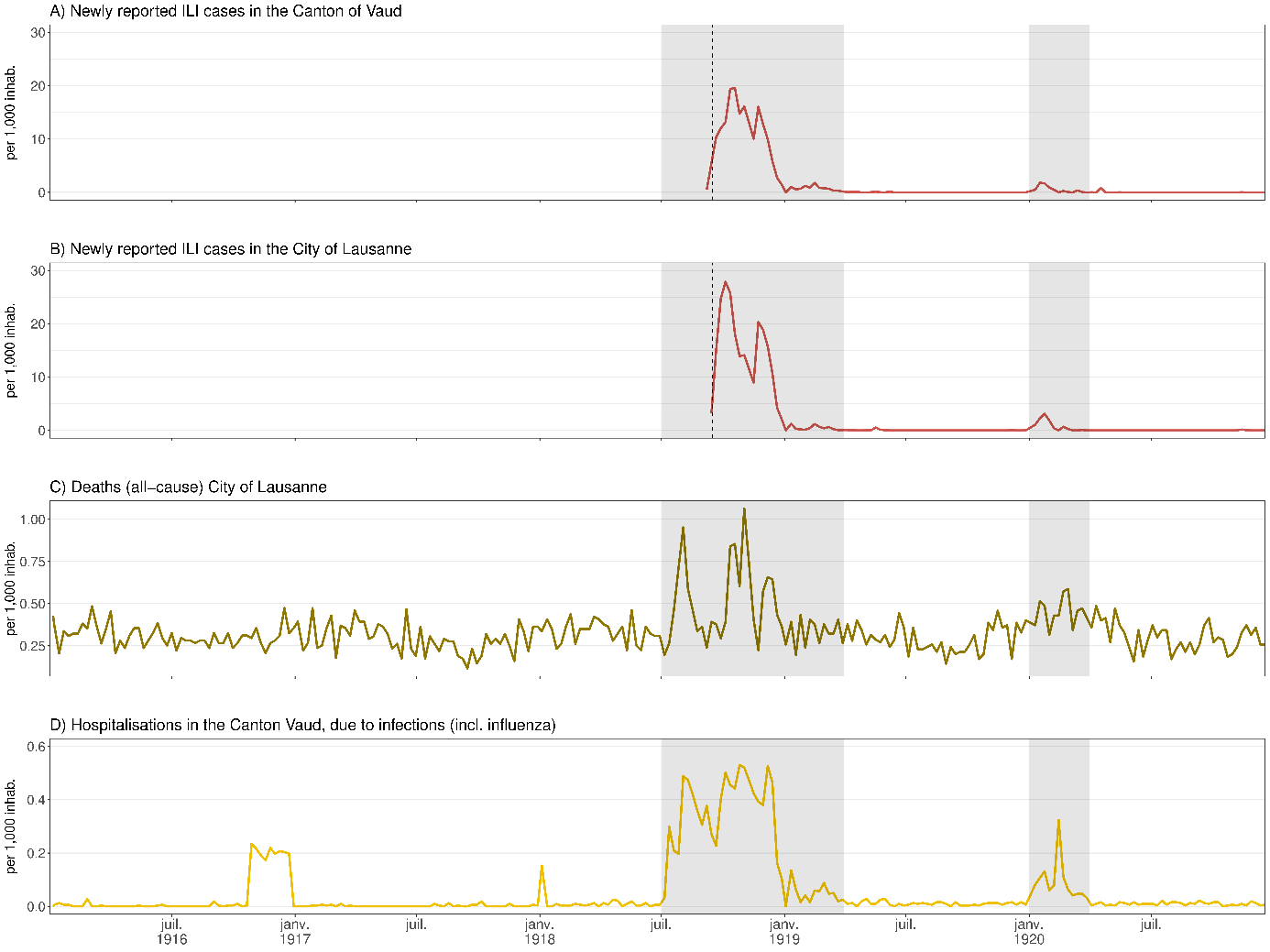

*Methods*

*Database and transcription:* a SQL database was designed and secured on the server of the University of Zurich, and an entry tool was coded on R using a ShinyApp. Quality controls were continuously performed.

*Variables*: civil status was categorized into “married” and “single or missing”, the latter grouping together “single”, “widowed”, “divorced” or missing. Syphilis status was based on symptoms or on the Wasserman test (Wassermann et al., 1906). Information on early neonatal mortality (first 7 days of life) is missing for mother-child pairs who left the maternity before 7 days post-partum. Hence, we used information concerning the first 5 days after delivery, since virtually all pairs stayed for that duration. Based on maternal residential address, a variable “living inside Lausanne” was defined, taking the values “yes”, “no”, or “unsure”. Since Lausanne residence was “unsure” in only 0.42% case, these were pooled with the category “no”. HISCO class is based on HISCO code and categorized as follows: 1 for unskilled workers/farm workers, 2 for medium-skilled workers, farmers, and 3 non-manual occupations, higher managers/professionals. Seasonality was categorized based on birth season (spring, summer, autumn, winter).

*ILI exposure variable:* infants *in utero* during the pandemic (based on its course in Lausanne, *Figure S1*, those born after 01/07/1918 and conceived before 01/04/1919, or born after 01/01/1920 and conceived before 01/04/1920), and whose mother had ILI during pregnancy, were considered as exposed to ILI (*n*=299). However, if the timing of the illness indicated that it occurred outside of the pandemic period, these cases were excluded (*n*=17), resulting in a total of *n*=282 infants exposed *in utero.* Regarding the timing of ILI, for women with two ILI episodes, the later date was used to estimate the trimester of disease onset. Regarding symptoms, they were considered severe if there was a mention of one of the following: bronchopneumonia, pneumonia, or bronchitis. In addition, a few (*n*=4) cases where it was clear the woman had developed severe symptoms were also qualified as such (“pulmonary complications, 3 weeks in bed”, “17 days of treatment at the hospital”, “very severe influenza with epistaxis”, “dry pleurisy”).

*Exclusion criteria*: birth weight, birth length, placenta weight and head circumference which had a sex-*specific Z*-score higher ≥ |7| standard deviations were excluded. This threshold is conservative because outliers are usually excluded at lower cut-offs (Phan et al., 2020).

*Statistical analyses:* multivariable GLMs were adjusted for variables (ILI, seasonality, maternal height, age and gravidity, neonatal sex), that we expected to be associated with the outcomes, based on the literature. Other potential explanatory variables (morphology, civil status, HISCO class, living inside Lausanne, goitre, rickets) were added using a stepwise algorithm. Based on the lowest Akaike information criterion (AIC), all models were adjusted for ILI, seasonality, maternal height, age, gravidity, neonatal sex, morphology, civil status, living inside Lausanne. GA is an important determinant of birth size (Wilcox & Skcerven, 1992), but it is also an intermediate variable on the causal path between exposure (disease) and outcome (neonatal size), since GA itself can be affected by maternal ILI and determines the time at which birth anthropometry is measured. Therefore, adjusting or stratifying for GA in the regression models may induce collider bias (Delbaere et al., 2007; Hernán et al., 2002; Skrivankova et al., 2023), thus we decided not to adjust our analyses on GA.

*R version and packages:* R version 4.4.1 was used (R Core Team, 2024). Tidyverse (Wickham et al., 2019) package was used for data processing and graphics.

**Figure S2: number of births per year**

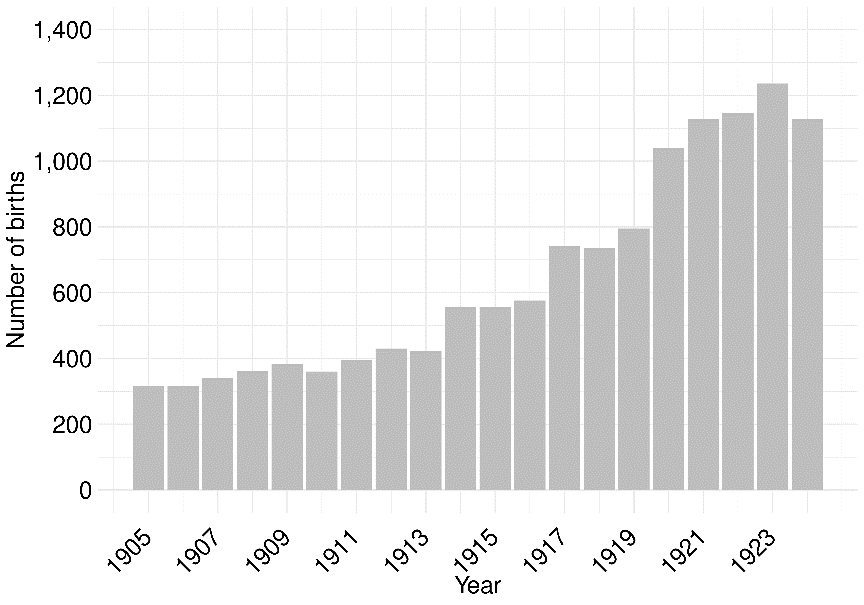

**Table S1: Maternal characteristics through the years (1905-1924)**

| **Variable** | **1905**, N = 315*^1^* | **1906**, N = 317*^1^* | **1907**, N = 340*^1^* | **1908**, N = 361*^1^* | **1909**, N = 383*^1^* | **1910**, N = 360*^1^* | **1911**, N = 396*^1^* | **1912**, N = 430*^1^* | **1913**, N = 423*^1^* | **1914**, N = 557*^1^* | **1915**, N = 557*^1^* | **1916**, N = 576*^1^* | **1917**, N = 741*^1^* | **1918**, N = 735*^1^* | **1919**, N = 795*^1^* | **1920**, N = 1,041*^1^* | **1921**, N = 1,129*^1^* | **1922**, N = 1,146*^1^* | **1923**, N = 1,236*^1^* | **1924**, N = 1,128*^1^* |
| --- | --- | --- | --- | --- | --- | --- | --- | --- | --- | --- | --- | --- | --- | --- | --- | --- | --- | --- | --- | --- |
| maternal age (years) | 27.8 (6.1) | 27.6 (6.7) | 27.7 (6.7) | 27.5 (6.4) | 28.2 (6.6) | 27.3 (6.4) | 28.3 (6.9) | 28.5 (6.8) | 27.7 (6.8) | 27.7 (6.5) | 28.2 (6.8) | 28.3 (6.4) | 28.2 (6.2) | 28.4 (6.5) | 28.6 (6.2) | 28.5 (6.2) | 28.3 (6.2) | 28.0 (5.9) | 28.4 (6.1) | 28.6 (6.0) |
| missing | 1 | 0 | 0 | 0 | 0 | 0 | 0 | 0 | 0 | 0 | 0 | 1 | 0 | 2 | 2 | 2 | 2 | 0 | 0 | 1 |
| height (cm) | 155.1 (5.9) | 154.3 (6.9) | 154.7 (6.6) | 155.1 (7.6) | 156.0 (6.5) | 155.9 (6.4) | 155.6 (6.9) | 155.7 (6.8) | 155.0 (6.1) | 154.9 (6.1) | 155.5 (6.4) | 155.8 (6.8) | 155.9 (6.6) | 156.0 (6.6) | 156.5 (6.1) | 156.3 (6.8) | 156.4 (6.6) | 156.5 (6.4) | 156.6 (6.0) | 156.7 (5.8) |
| missing | 16 | 13 | 7 | 7 | 22 | 9 | 12 | 16 | 24 | 23 | 15 | 8 | 20 | 31 | 36 | 47 | 45 | 38 | 57 | 50 |
| civil status |  |  |  |  |  |  |  |  |  |  |  |  |  |  |  |  |  |  |  |  |
| married | 233 (74%) | 208 (66%) | 217 (64%) | 260 (72%) | 277 (72%) | 256 (71%) | 292 (74%) | 324 (75%) | 319 (75%) | 424 (76%) | 445 (80%) | 489 (85%) | 616 (83%) | 619 (84%) | 694 (87%) | 925 (89%) | 1,010 (89%) | 1,052 (92%) | 1,133 (92%) | 1,043 (92%) |
| single or missing | 82 (26%) | 109 (34%) | 123 (36%) | 101 (28%) | 106 (28%) | 104 (29%) | 104 (26%) | 106 (25%) | 104 (25%) | 133 (24%) | 112 (20%) | 87 (15%) | 125 (17%) | 116 (16%) | 101 (13%) | 116 (11%) | 119 (11%) | 94 (8%) | 103 (8%) | 85 (8%) |
| gravidity |  |  |  |  |  |  |  |  |  |  |  |  |  |  |  |  |  |  |  |  |
| 1 | 94 (30%) | 122 (38%) | 119 (35%) | 126 (35%) | 118 (31%) | 131 (36%) | 135 (34%) | 143 (33%) | 158 (37%) | 191 (34%) | 182 (33%) | 186 (32%) | 283 (38%) | 290 (39%) | 284 (36%) | 447 (43%) | 473 (42%) | 430 (38%) | 454 (37%) | 422 (37%) |
| 2 | 63 (20%) | 65 (21%) | 69 (20%) | 77 (21%) | 81 (21%) | 88 (24%) | 74 (19%) | 92 (21%) | 89 (21%) | 116 (21%) | 137 (25%) | 109 (19%) | 151 (20%) | 143 (19%) | 205 (26%) | 209 (20%) | 247 (22%) | 281 (25%) | 287 (23%) | 257 (23%) |
| >2 | 158 (50%) | 130 (41%) | 152 (45%) | 158 (44%) | 184 (48%) | 141 (39%) | 187 (47%) | 195 (45%) | 176 (42%) | 250 (45%) | 236 (43%) | 281 (49%) | 307 (41%) | 302 (41%) | 306 (38%) | 385 (37%) | 409 (36%) | 435 (38%) | 495 (40%) | 448 (40%) |
| missing | 0 | 0 | 0 | 0 | 0 | 0 | 0 | 0 | 0 | 0 | 2 | 0 | 0 | 0 | 0 | 0 | 0 | 0 | 0 | 1 |
| living in Lausanne | 158 (50%) | 166 (52%) | 175 (51%) | 179 (50%) | 191 (50%) | 186 (52%) | 203 (51%) | 239 (56%) | 217 (51%) | 287 (52%) | 281 (50%) | 265 (46%) | 344 (46%) | 364 (50%) | 360 (45%) | 454 (44%) | 453 (40%) | 437 (38%) | 446 (36%) | 422 (37%) |
| morphology |  |  |  |  |  |  |  |  |  |  |  |  |  |  |  |  |  |  |  |  |
| neither | 309 (98%) | 297 (94%) | 337 (99%) | 356 (99%) | 381 (99%) | 358 (99%) | 383 (97%) | 405 (94%) | 383 (91%) | 498 (89%) | 487 (87%) | 541 (94%) | 702 (95%) | 699 (95%) | 743 (93%) | 970 (93%) | 1,077 (95%) | 1,080 (94%) | 1,206 (98%) | 1,043 (92%) |
| obese | 1 (0%) | 9 (3%) | 2 (1%) | 4 (1%) | 2 (1%) | 1 (0%) | 5 (1%) | 17 (4%) | 25 (6%) | 33 (6%) | 33 (6%) | 16 (3%) | 11 (1%) | 19 (3%) | 27 (3%) | 44 (4%) | 37 (3%) | 43 (4%) | 22 (2%) | 59 (5%) |
| thin | 5 (2%) | 11 (3%) | 1 (0%) | 1 (0%) | 0 (0%) | 1 (0%) | 8 (2%) | 8 (2%) | 15 (4%) | 26 (5%) | 37 (7%) | 19 (3%) | 28 (4%) | 17 (2%) | 25 (3%) | 27 (3%) | 15 (1%) | 23 (2%) | 8 (1%) | 26 (2%) |
| goitre | 14 (4%) | 33 (10%) | 19 (6%) | 23 (6%) | 42 (11%) | 27 (8%) | 21 (5%) | 26 (6%) | 31 (7%) | 58 (10%) | 97 (17%) | 55 (10%) | 26 (4%) | 20 (3%) | 45 (6%) | 72 (7%) | 98 (9%) | 47 (4%) | 17 (1%) | 7 (1%) |
| rickets | 10 (3%) | 43 (14%) | 18 (5%) | 44 (12%) | 15 (4%) | 10 (3%) | 37 (9%) | 57 (13%) | 50 (12%) | 104 (19%) | 125 (22%) | 69 (12%) | 40 (5%) | 44 (6%) | 35 (4%) | 42 (4%) | 30 (3%) | 25 (2%) | 8 (1%) | 15 (1%) |
| flu during pregnancy and pandemic | 0 (0%) | 0 (0%) | 0 (0%) | 0 (0%) | 0 (0%) | 0 (0%) | 0 (0%) | 0 (0%) | 0 (0%) | 0 (0%) | 0 (0%) | 0 (0%) | 0 (0%) | 57 (8%) | 155 (19%) | 70 (7%) | 0 (0%) | 0 (0%) | 0 (0%) | 0 (0%) |
| syphilis | 5 (2%) | 4 (1%) | 3 (1%) | 5 (1%) | 8 (2%) | 16 (4%) | 7 (2%) | 6 (1%) | 3 (1%) | 8 (1%) | 10 (2%) | 12 (2%) | 6 (1%) | 6 (1%) | 8 (1%) | 13 (1%) | 18 (2%) | 7 (1%) | 15 (1%) | 7 (1%) |
| maternal mortality | 2 (1%) | 4 (1%) | 2 (1%) | 4 (1%) | 2 (1%) | 3 (1%) | 2 (1%) | 4 (1%) | 2 (0%) | 9 (2%) | 7 (1%) | 6 (1%) | 2 (0%) | 6 (1%) | 0 (0%) | 4 (0%) | 6 (1%) | 2 (0%) | 7 (1%) | 2 (0%) |
| HISCO class |  |  |  |  |  |  |  |  |  |  |  |  |  |  |  |  |  |  |  |  |
| 2 | 218 (69%) | 210 (66%) | 246 (72%) | 276 (76%) | 305 (80%) | 269 (75%) | 291 (73%) | 333 (77%) | 324 (77%) | 434 (78%) | 444 (80%) | 525 (91%) | 640 (86%) | 632 (86%) | 684 (86%) | 894 (86%) | 995 (88%) | 1,067 (93%) | 1,144 (93%) | 472 (42%) |
| 1 | 76 (24%) | 90 (28%) | 73 (21%) | 62 (17%) | 58 (15%) | 70 (19%) | 76 (19%) | 70 (16%) | 71 (17%) | 92 (17%) | 84 (15%) | 38 (7%) | 55 (7%) | 47 (6%) | 53 (7%) | 57 (5%) | 60 (5%) | 37 (3%) | 50 (4%) | 59 (5%) |
| 3 | 4 (1%) | 5 (2%) | 6 (2%) | 11 (3%) | 6 (2%) | 4 (1%) | 6 (2%) | 12 (3%) | 9 (2%) | 14 (3%) | 13 (2%) | 4 (1%) | 12 (2%) | 18 (2%) | 17 (2%) | 15 (1%) | 15 (1%) | 8 (1%) | 7 (1%) | 15 (1%) |
| missing | 17 (5%) | 12 (4%) | 15 (4%) | 12 (3%) | 14 (4%) | 17 (5%) | 23 (6%) | 15 (3%) | 19 (4%) | 17 (3%) | 16 (3%) | 9 (2%) | 34 (5%) | 38 (5%) | 41 (5%) | 75 (7%) | 59 (5%) | 34 (3%) | 35 (3%) | 582 (52%) |
| season |  |  |  |  |  |  |  |  |  |  |  |  |  |  |  |  |  |  |  |  |
| spring | 72 (23%) | 106 (33%) | 91 (27%) | 98 (27%) | 100 (26%) | 82 (23%) | 119 (30%) | 119 (28%) | 113 (27%) | 164 (29%) | 169 (30%) | 141 (24%) | 188 (25%) | 206 (28%) | 181 (23%) | 265 (25%) | 300 (27%) | 315 (27%) | 335 (27%) | 307 (27%) |
| summer | 75 (24%) | 72 (23%) | 92 (27%) | 98 (27%) | 106 (28%) | 103 (29%) | 84 (21%) | 117 (27%) | 107 (25%) | 143 (26%) | 129 (23%) | 153 (27%) | 204 (28%) | 185 (25%) | 187 (24%) | 260 (25%) | 298 (26%) | 301 (26%) | 295 (24%) | 274 (24%) |
| autumn | 91 (29%) | 82 (26%) | 69 (20%) | 84 (23%) | 96 (25%) | 88 (24%) | 87 (22%) | 100 (23%) | 110 (26%) | 118 (21%) | 102 (18%) | 137 (24%) | 179 (24%) | 176 (24%) | 241 (30%) | 251 (24%) | 261 (23%) | 267 (23%) | 300 (24%) | 261 (23%) |
| winter | 77 (24%) | 57 (18%) | 88 (26%) | 81 (22%) | 81 (21%) | 87 (24%) | 106 (27%) | 94 (22%) | 93 (22%) | 132 (24%) | 157 (28%) | 145 (25%) | 170 (23%) | 168 (23%) | 186 (23%) | 265 (25%) | 270 (24%) | 263 (23%) | 306 (25%) | 286 (25%) |
| *^1^*Mean (SD); n (%) | | | | | | | | | | | | | | | | | | | | |

**Table S2: Neonatal characteristics through the years (1905-1924)**

| **Variable** | **1905**, N = 315*^1^* | **1906**, N = 317*^1^* | **1907**, N = 340*^1^* | **1908**, N = 361*^1^* | **1909**, N = 383*^1^* | **1910**, N = 360*^1^* | **1911**, N = 396*^1^* | **1912**, N = 430*^1^* | **1913**, N = 423*^1^* | **1914**, N = 557*^1^* | **1915**, N = 557*^1^* | **1916**, N = 576*^1^* | **1917**, N = 741*^1^* | **1918**, N = 735*^1^* | **1919**, N = 795*^1^* | **1920**, N = 1,041*^1^* | **1921**, N = 1,129*^1^* | **1922**, N = 1,146*^1^* | **1923**, N = 1,236*^1^* | **1924**, N = 1,128*^1^* |
| --- | --- | --- | --- | --- | --- | --- | --- | --- | --- | --- | --- | --- | --- | --- | --- | --- | --- | --- | --- | --- |
| birth weight (g) | 3,042.9 (602.5) | 3,104.3 (571.4) | 3,085.9 (560.4) | 3,039.4 (562.9) | 3,089.7 (604.0) | 3,048.7 (601.7) | 3,093.5 (607.5) | 3,050.7 (580.7) | 3,095.5 (600.4) | 3,201.5 (572.9) | 3,215.2 (554.4) | 3,177.6 (549.0) | 3,156.0 (547.0) | 3,132.1 (567.1) | 3,168.7 (588.2) | 3,198.9 (567.9) | 3,209.5 (540.7) | 3,221.9 (503.5) | 3,220.3 (544.4) | 3,294.0 (520.0) |
| head circumference (cm) | 33.6 (1.7) | 33.6 (1.7) | 34.0 (1.8) | 34.5 (1.7) | 34.8 (2.2) | 33.9 (2.2) | 34.3 (1.8) | 34.1 (1.9) | 34.0 (2.1) | 34.3 (2.0) | 34.4 (1.9) | 34.5 (1.8) | 34.3 (1.7) | 34.4 (1.9) | 34.4 (1.9) | 34.3 (1.8) | 34.5 (1.7) | 34.5 (1.5) | 34.5 (1.8) | 34.7 (1.6) |
| missing | 12 | 11 | 3 | 12 | 9 | 8 | 10 | 5 | 8 | 8 | 5 | 7 | 9 | 12 | 9 | 25 | 12 | 26 | 13 | 9 |
| placenta weight (g) | 568.3 (130.6) | 553.3 (107.2) | 557.5 (130.7) | 571.6 (123.2) | 575.6 (125.0) | 566.1 (122.8) | 559.1 (124.4) | 544.3 (114.1) | 551.2 (122.4) | 569.4 (122.2) | 558.9 (117.8) | 575.9 (124.9) | 572.9 (128.3) | 570.2 (125.6) | 584.2 (119.2) | 580.7 (117.0) | 590.2 (126.8) | 591.1 (118.4) | 584.1 (124.8) | 579.3 (113.0) |
| missing | 2 | 3 | 1 | 0 | 3 | 1 | 3 | 4 | 3 | 8 | 6 | 10 | 9 | 17 | 7 | 18 | 25 | 13 | 7 | 5 |
| birth length (cm) | 48.9 (3.3) | 48.7 (2.6) | 48.7 (2.5) | 48.5 (2.9) | 48.8 (3.2) | 48.6 (3.1) | 49.2 (3.0) | 48.6 (2.8) | 49.1 (3.1) | 49.4 (2.7) | 49.3 (2.8) | 49.2 (2.7) | 49.1 (2.3) | 49.0 (2.9) | 49.0 (2.9) | 49.0 (2.7) | 49.2 (2.5) | 49.3 (2.4) | 49.3 (2.5) | 49.6 (2.2) |
| missing | 2 | 1 | 1 | 1 | 1 | 1 | 1 | 0 | 4 | 1 | 2 | 0 | 5 | 3 | 2 | 3 | 5 | 1 | 2 | 2 |
| ponderal index | 2.6 (0.3) | 2.7 (0.3) | 2.6 (0.3) | 2.7 (0.3) | 2.6 (0.3) | 2.6 (0.3) | 2.6 (0.3) | 2.6 (0.3) | 2.6 (0.3) | 2.6 (0.2) | 2.7 (0.3) | 2.6 (0.3) | 2.7 (0.3) | 2.6 (0.3) | 2.7 (0.3) | 2.7 (0.3) | 2.7 (0.3) | 2.7 (0.3) | 2.7 (0.3) | 2.7 (0.2) |
| missing | 2 | 1 | 1 | 1 | 1 | 1 | 1 | 0 | 4 | 1 | 2 | 0 | 5 | 3 | 2 | 3 | 5 | 1 | 2 | 2 |
| gestational age (weeks) | 38.9 (2.4) | 39.0 (2.1) | 39.1 (2.1) | 38.9 (2.3) | 38.9 (2.5) | 38.7 (2.5) | 38.9 (2.3) | 38.8 (2.5) | 39.0 (2.4) | 39.3 (2.2) | 39.3 (2.2) | 38.9 (2.2) | 39.0 (2.0) | 39.2 (2.3) | 39.1 (2.2) | 39.2 (2.1) | 39.3 (2.0) | 39.5 (1.7) | 39.4 (1.8) | 39.7 (1.7) |
| sex |  |  |  |  |  |  |  |  |  |  |  |  |  |  |  |  |  |  |  |  |
| Male | 155 (49%) | 165 (52%) | 179 (53%) | 177 (49%) | 183 (48%) | 197 (55%) | 199 (51%) | 222 (52%) | 210 (50%) | 293 (53%) | 274 (49%) | 312 (54%) | 388 (52%) | 383 (52%) | 402 (51%) | 537 (52%) | 573 (51%) | 558 (49%) | 658 (53%) | 556 (49%) |
| Female | 159 (51%) | 152 (48%) | 160 (47%) | 184 (51%) | 200 (52%) | 163 (45%) | 195 (49%) | 208 (48%) | 213 (50%) | 261 (47%) | 282 (51%) | 264 (46%) | 353 (48%) | 352 (48%) | 391 (49%) | 501 (48%) | 546 (49%) | 585 (51%) | 577 (47%) | 570 (51%) |
| missing | 1 | 0 | 1 | 0 | 0 | 0 | 2 | 0 | 0 | 3 | 1 | 0 | 0 | 0 | 2 | 3 | 10 | 3 | 1 | 2 |
| low birth weight (<2'500g) | 44 (14%) | 39 (12%) | 40 (12%) | 46 (13%) | 51 (13%) | 46 (13%) | 50 (13%) | 63 (15%) | 50 (12%) | 51 (9%) | 42 (8%) | 52 (9%) | 60 (8%) | 72 (10%) | 81 (10%) | 87 (8%) | 89 (8%) | 67 (6%) | 92 (7%) | 64 (6%) |
| preterm birth (<37 weeks) | 32 (10%) | 39 (12%) | 38 (11%) | 35 (10%) | 43 (11%) | 42 (12%) | 46 (12%) | 68 (16%) | 41 (10%) | 49 (9%) | 52 (9%) | 83 (14%) | 100 (13%) | 69 (9%) | 68 (9%) | 90 (9%) | 82 (7%) | 70 (6%) | 74 (6%) | 53 (5%) |
| stillbirth | 20 (6%) | 19 (6%) | 18 (5%) | 21 (6%) | 23 (6%) | 17 (5%) | 21 (5%) | 15 (3%) | 18 (4%) | 20 (4%) | 20 (4%) | 23 (4%) | 25 (3%) | 36 (5%) | 32 (4%) | 42 (4%) | 27 (2%) | 36 (3%) | 47 (4%) | 36 (3%) |
| neonatal mortality d1-5 | 5 (2%) | 12 (4%) | 10 (3%) | 8 (2%) | 18 (5%) | 15 (4%) | 20 (5%) | 19 (4%) | 14 (3%) | 12 (2%) | 12 (2%) | 13 (2%) | 17 (2%) | 14 (2%) | 22 (3%) | 18 (2%) | 26 (2%) | 13 (1%) | 16 (1%) | 21 (2%) |
| missing | 1 | 1 | 0 | 0 | 3 | 1 | 0 | 0 | 1 | 1 | 0 | 0 | 0 | 2 | 0 | 0 | 2 | 1 | 0 | 0 |
| *^1^*Mean (SD); n (%) | | | | | | | | | | | | | | | | | | | | |

**Figure S3:** annual maternal characteristics of the population, 1905-1924

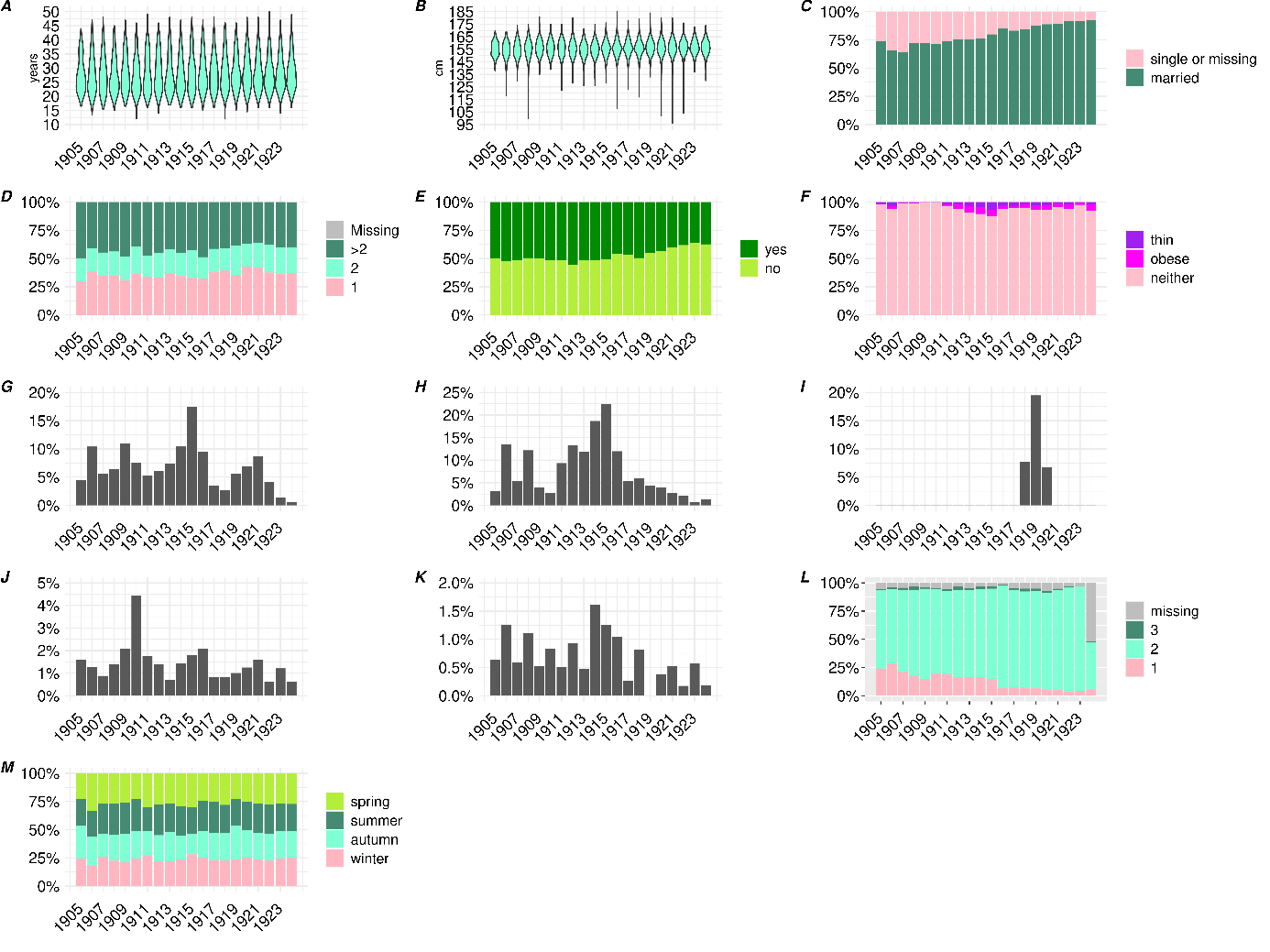

Maternal age (**A**), height (**B**), civil status region (**C**), gravidity (**D**), living in the city of Lausanne at the time of delivery (**E**), morphology (**F**), goitre (**G**), rickets (**H**), influenza during pregnancy and during the pandemic (**I**), syphilis diagnosis (**J**), maternal mortality (**K**), HISCO class (**L**), season (**M**)

**Figure S4:** annual neonatal characteristics of the population, 1905-1924

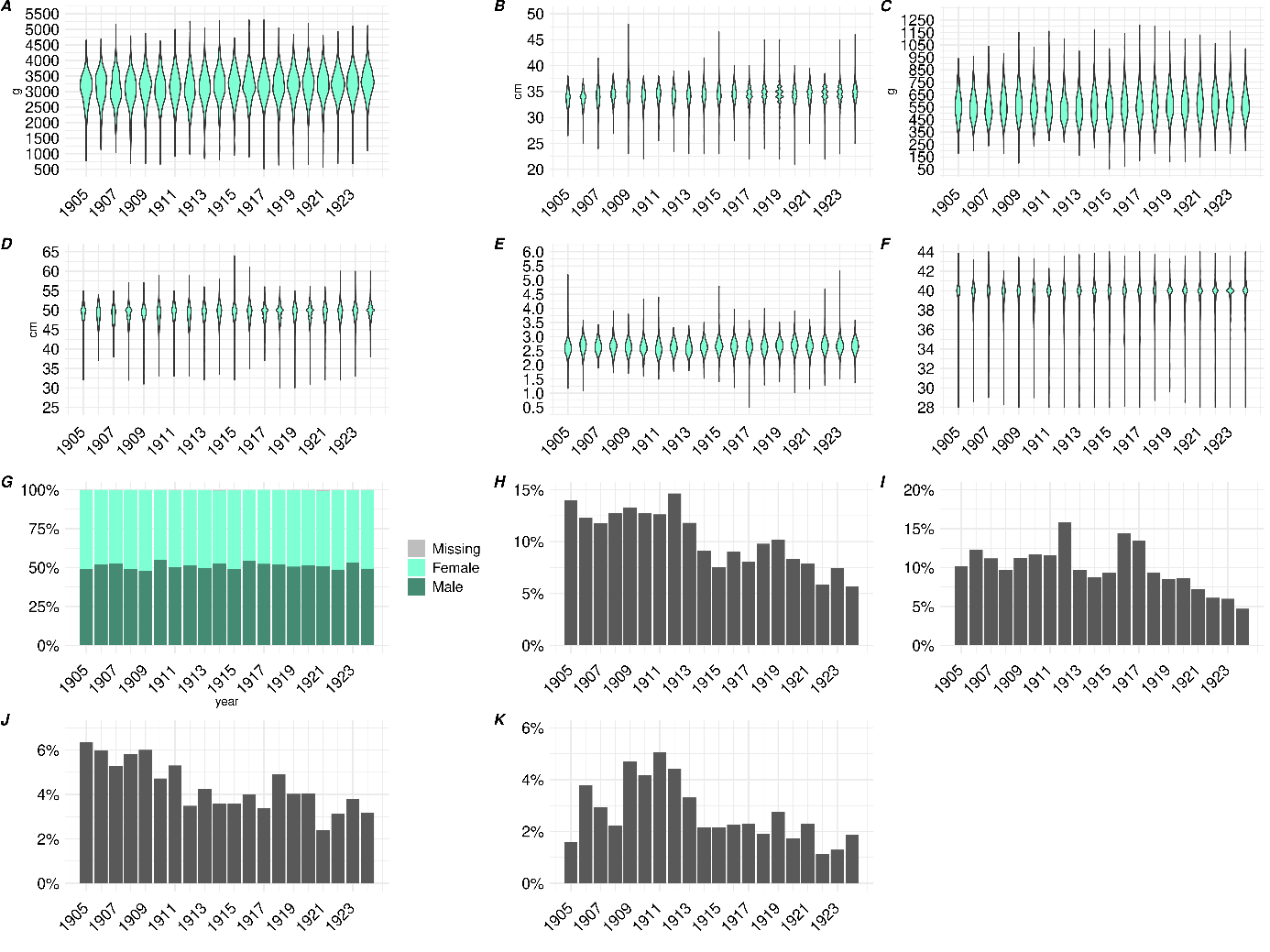

Birth weight (**A**), head circumference (**B**), placenta weight (**C**), birth length (**D**), ponderal index (**E**), gestational age (**F**), sex (**G**), low birth weight (<2’500g, **H**), preterm birth (<37 weeks, **I**), stillbirth (**J**), early neonatal mortality in the first five days after birth (**K**)

**Table S3: Maternal and neonatal characteristics during the pandemic.** *^1^*Mean (SD); n (%).

| **Maternal characteristics** | | season |  |
| --- | --- | --- | --- |
| **Variable** | ***n* = 2,177***^1^* | spring | 446 (20%) |
| maternal age (years) | 28.5 (6.2) | summer | 569 (26%) |
| missing | 4 | autumn | 666 (31%) |
| height (cm) | 156.4 (6.5) | winter | 496 (23%) |
| missing | 101 | maternal mortality | 10 (0%) |
| civil status |  | **Neonatal characteristics** | |
| married | 1,918 (88%) | **Variable** | ***n* =2,177*^1^*** |
| single or missing | 259 (12%) | birth weight (g) | 3,187.8 (566.9) |
| gravidity |  | head circumference (cm) | 34.4 (1.8) |
| 1 | 4,788 (37%) | missing | 37 |
| 2 | 2,840 (22%) | placenta weight (g) | 581.9 (118.4) |
| >2 | 5,335 (41%) | missing | 29 |
| living in Lausanne | 981 (45%) | birth length (cm) | 49.1 (2.7) |
| morphology |  | missing | 6 |
| neither | 2,031 (93%) | ponderal index | 2.7 (0.3) |
| obese | 85 (4%) | missing | 6 |
| thin | 61 (3%) | gestational age (weeks) | 39.2 (2.1) |
| goitre | 122 (6%) | sex |  |
| rickets | 87 (4%) | Male | 1,122 (52%) |
| flu during pregnancy and pandemic | 282 (13%) | Female | 1,050 (48%) |
| syphilis | 24 (1%) | missing | 5 |
| HISCO class |  | low birth weight (<2'500g) | 195 (9%) |
| 2 | 1,876 (86%) | preterm birth (<37 weeks) | 186 (9%) |
| 1 | 127 (6%) | stillbirth | 92 (4%) |
| 3 | 39 (2%) | neonatal mortality d1-5 | 43 (2%) |
| missing | 135 (6%) | missing | 1 |

**DIRECT EXPOSURE TO THE INFLUENZA PANDEMIC**

**Table S4: Maternal characteristics depending on influenza during the pregnancy (during the pandemic period only).** p: p-value. *^1^*Mean (SD); n (%). *^2^*Wilcoxon rank sum test; Pearson's Chi-squared test; Fisher's exact test

| **ILI** | **no**, *n* = 1,895*^1^* | **yes**, *n* = 282*^1^* | **p-value***^2^* |
| --- | --- | --- | --- |
| maternal age (years) | 28.5 (6.2) | 28.6 (6.2) | 0.8 |
| missing | 4 | 0 |  |
| height (cm) | 156.4 (6.5) | 156.1 (6.8) | 0.3 |
| missing | 90 | 11 |  |
| civil status |  |  | 0.8 |
| married | 1,668 (88%) | 250 (89%) |  |
| single or missing | 227 (12%) | 32 (11%) |  |
| gravidity |  |  | 0.4 |
| 1 | 758 (40%) | 104 (37%) |  |
| 2 | 413 (22%) | 70 (25%) |  |
| >2 | 724 (38%) | 108 (38%) |  |
| living in Lausanne | 851 (45%) | 130 (46%) | 0.7 |
| morphology |  |  | 0.8 |
| neither | 1,770 (93%) | 261 (93%) |  |
| obese | 72 (4%) | 13 (5%) |  |
| thin | 53 (3%) | 8 (3%) |  |
| goitre | 106 (6%) | 16 (6%) | >0.9 |
| rickets | 76 (4%) | 11 (4%) | >0.9 |
| syphilis | 20 (1%) | 4 (1%) | 0.5 |
| maternal mortality | 7 (0%) | 3 (1%) | 0.13 |
| HISCO class |  |  | 0.7 |
| 2 | 1,627 (86%) | 249 (88%) |  |
| 1 | 112 (6%) | 15 (5%) |  |
| 3 | 34 (2%) | 5 (2%) |  |
| missing | 122 (6%) | 13 (5%) |  |
| season |  |  | <0.001 |
| spring | 344 (18%) | 102 (36%) |  |
| summer | 502 (26%) | 67 (24%) |  |
| autumn | 628 (33%) | 38 (13%) |  |
| winter | 421 (22%) | 75 (27%) |  |

**Figure S5: counts and dates of *n*=227 reported influenza-like-illness during the pandemic (note: the exact month was unknown for *n*=55 cases of the 282 ILI cases).**

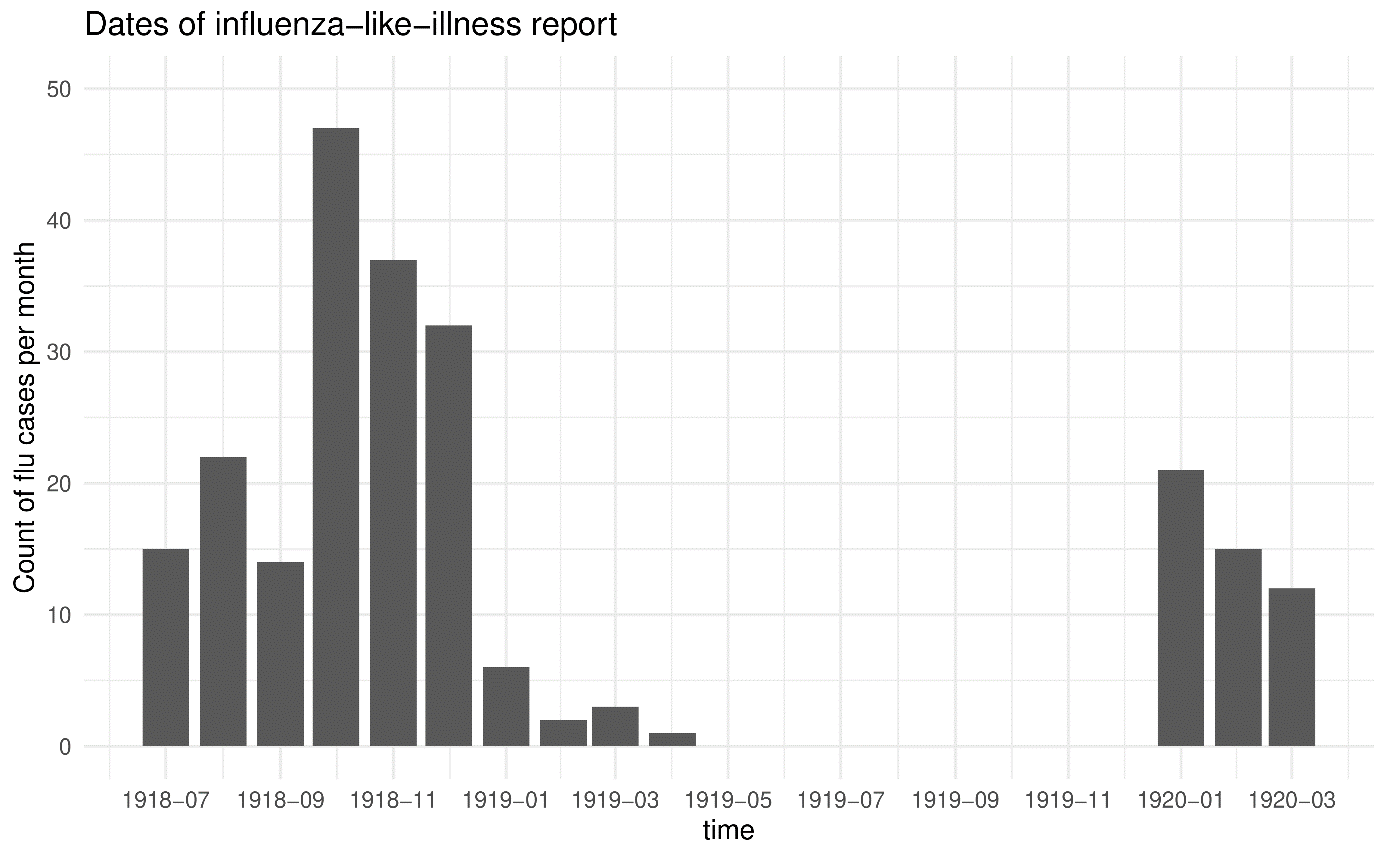

**Table S5: pregnancy outcomes depending on maternal ILI, univariable and multivariable GLMs, all births and stratified by sex (only using livebirths except for stillbirth outcome).** Full tables for the multivariable models among all births (males and females) are displayed in *Tables S5-7*. p: p-value. *^1^*adjusted for: ILI during pregnancy, maternal age, maternal height, gravidity, sex (except for the sex-stratified models), season, morphology, civil status, Lausanne.

|  |  | Univariable | | | | | Multivariable*^1^* | | | | |
| --- | --- | --- | --- | --- | --- | --- | --- | --- | --- | --- | --- |
|  |  |  | 95% CI | |  | |  | 95% CI | |  | |
|  | Births | beta | lci | uci | p | *n* | beta | lci | uci | p | *n* |
| Birth weight (g) | all | **-120.01** | **-187.36** | **-52.65** | **<0.0001** | 2,085 | **-93.62** | **-159.89** | **-27.35** | **0.01** | 1,999 |
|  | males | **-137.56** | **-241.45** | **-33.67** | **0.01** | 1,069 |  |  | - |  |  |
|  | females | **-89.44** | **-175.54** | **-3.34** | **0.04** | 1,013 |  |  | - |  |  |
| Head circumference (cm) | all | **-0.26** | **-0.48** | **-0.05** | **0.01** | 2,077 | -0.14 | -0.35 | 0.07 | 0.18 | 1,992 |
|  | males | **-0.43** | **-0.74** | **-0.11** | **0.01** | 1,066 |  |  | - |  |  |
|  | females | -0.04 | -0.31 | 0.24 | 0.78 | 1,008 |  |  | - |  |  |
| Placenta weight (g) | all | **-36.65** | **-51.29** | **-22.02** | **<0.0001** | -36.65 | **-30.73** | **-45.49** | **-15.97** | **<0.0001** | 1,977 |
|  | males | **-44.92** | **-67.30** | **-22.53** | **<0.0001** | 1,053 |  |  | - |  |  |
|  | females | **-27.19** | **-46.20** | **-8.19** | **0.01** | 1,003 |  |  | - |  |  |
| Birth length (cm) | all | **-0.42** | **-0.73** | **-0.11** | **0.01** | 2,081 | **-0.32** | **-0.63** | **-0.01** | **0.04** | 1,997 |
|  | males | **-0.47** | **-0.93** | **0.00** | **0.05** | 1,068 |  |  | - |  |  |
|  | females | -0.31 | -0.72 | 0.10 | 0.14 | 1,010 |  |  | - |  |  |
| Ponderal index | all | **-0.04** | **-0.07** | **-0.01** | **0.02** | 2,081 | **-0.03** | **-0.06** | **0.00** | **0.04** | 1,997 |
|  | males | **-0.05** | **-0.09** | **0.00** | **0.05** | 1,068 |  |  | **-** |  |  |
|  | females | -0.03 | -0.07 | 0.01 | 0.19 | 1,010 |  |  | - |  |  |
| Gestational age (weeks) | all | -0.23 | -0.47 | 0.01 | 0.06 | 2,085 | -0.19 | -0.43 | 0.05 | 0.13 | 1,999 |
|  | males | -0.22 | -0.58 | 0.13 | 0.22 | 1,069 |  |  | - |  |  |
|  | females | -0.23 | -0.55 | 0.09 | 0.16 | 1,013 |  |  | - |  |  |
|  |  |  | 95% CI | |  | |  | 95% CI | |  | |
|  |  | OR | lci | uci | p-value | n | OR | lci | uci | p-value | n |
| Low birth weight (<2’500g) | all | **1.82** | **1.20** | **2.74** | **<0.0001** | 2,085 | **2.06** | **1.33** | **3.20** | **<0.0001** | 1,999 |
|  | males | **1.92** | **1.08** | **3.43** | **0.03** | 1,069 |  |  | - |  |  |
|  | females | 1.75 | 0.98 | 3.14 | 0.06 | 1,013 |  |  | - |  |  |
| Preterm birth (<37 weeks) | all | **1.61** | **1.04** | **2.47** | **0.03** | 2,085 | 1.55 | 0.97 | 2.46 | 0.07 | 1,999 |
|  | males | 1.67 | 0.89 | 3.14 | 0.11 | 1,069 |  |  | - |  |  |
|  | females | 1.54 | 0.85 | 2.80 | 0.15 | 1,013 |  |  | - |  |  |
| Stillbirth | all | 1.01 | 0.54 | 1.87 | 0.98 | 2,177 | - | | | | |
|  | males | 1.38 | 0.64 | 3.00 | 0.42 | 1,122 |  |  |  |  |  |
|  | females | 0.71 | 0.25 | 2.03 | 0.52 | 1,050 |  |  |  |  |  |
| Neonatal mortality day 1-5 | all | 1.10 | 0.46 | 2.62 | 0.84 | 2,084 | - | | | | |
|  | males | 0.91 | 0.21 | 3.99 | 0.90 | 1,069 |  |  |  |  |  |
|  | females | 1.18 | 0.40 | 3.51 | 0.76 | 1,012 |  |  |  |  |  |

**Table S6: pregnancy outcomes depending on maternal ILI, multivariable GLMs for birth weight, head circumference and placenta weight (only using livebirths), full tables.** p: p-value.

|  |  | **Birth weight (g)** | | | | **Head circumference (cm)** | | | | **Placenta weight (g)** | | | |
| --- | --- | --- | --- | --- | --- | --- | --- | --- | --- | --- | --- | --- | --- |
|  |  |  | 95% CI | |  |  | 95% CI | |  |  | 95% CI | |  |
| Parameters | category | beta | lci | uci | pvalue | beta | lci | uci | pvalue | beta | lci | uci | pvalue |
| Influenza during pregnancy (ref: no) | yes | -93.62 | -159.89 | -27.35 | 0.01 | -0.14 | -0.35 | 0.07 | 0.18 | -30.73 | -45.49 | -15.97 | <0.0001 |
| Maternal age |  | -0.92 | -5.26 | 3.41 | 0.68 | 0.00 | -0.01 | 0.02 | 0.69 | 0.97 | 0.00 | 1.94 | 0.05 |
| Maternal height |  | 14.53 | 11.14 | 17.92 | <0.0001 | 0.03 | 0.02 | 0.04 | <0.0001 | 2.50 | 1.74 | 3.26 | <0.0001 |
| Gravidity (ref: 1) | 2 | 106.04 | 45.98 | 166.09 | <0.0001 | 0.09 | -0.10 | 0.28 | 0.37 | 13.57 | 0.17 | 26.98 | 0.05 |
|  | >2 | 187.33 | 126.70 | 247.95 | <0.0001 | 0.00 | -0.19 | 0.19 | 0.98 | 13.84 | 0.26 | 27.41 | 0.05 |
| Sex (ref: male) | female | -117.87 | -161.85 | -73.90 | <0.0001 | -0.67 | -0.81 | -0.53 | <0.0001 | -20.01 | -29.83 | -10.19 | <0.0001 |
| Season (ref: spring) | summer | 20.49 | -44.42 | 85.40 | 0.54 | 0.15 | -0.06 | 0.35 | 0.16 | 7.89 | -6.60 | 22.39 | 0.29 |
|  | autumn | 56.77 | -6.98 | 120.53 | 0.08 | 0.24 | 0.04 | 0.44 | 0.02 | 20.22 | 5.98 | 34.47 | 0.01 |
|  | winter | 67.74 | 1.13 | 134.34 | 0.05 | 0.26 | 0.05 | 0.47 | 0.02 | -0.67 | -15.52 | 14.19 | 0.93 |
| Morphology (ref: none) | obese | 180.62 | 66.45 | 294.79 | <0.0001 | 0.56 | 0.20 | 0.92 | <0.0001 | 47.14 | 21.60 | 72.68 | <0.0001 |
|  | thin | -295.94 | -430.23 | -161.64 | <0.0001 | -0.80 | -1.22 | -0.37 | <0.0001 | -47.40 | -77.25 | -17.56 | <0.0001 |
| Civil status (ref: married) | single or missing | -84.10 | -154.69 | -13.51 | 0.02 | -0.34 | -0.56 | -0.12 | <0.0001 | -13.44 | -29.26 | 2.38 | 0.10 |
| Living inside Lausanne (ref: yes) | no or unsure | 80.23 | 36.08 | 124.38 | <0.0001 | 0.35 | 0.21 | 0.49 | <0.0001 | 18.31 | 8.45 | 28.18 | <0.0001 |
| *n* |  | 1,999 | | | | 1,992 | | | | 1,977 | | | |

**Table S7: pregnancy outcomes depending on maternal ILI, multivariable GLMs for birth length, ponderal index and gestational age (only using livebirths), full tables.** p: p-value.

|  |  | **Birth length (cm)** | | | | **Ponderal index** | | | | **Gestational age (weeks)** | | | |
| --- | --- | --- | --- | --- | --- | --- | --- | --- | --- | --- | --- | --- | --- |
|  |  |  | 95% CI | |  |  | 95% CI | |  |  | 95% CI | |  |
| Parameters | category | beta | lci | uci | pvalue | beta | lci | uci | pvalue | beta | lci | uci | pvalue |
| Influenza during pregnancy (ref: no) | yes | -0.32 | -0.63 | -0.01 | 0.04 | -0.03 | -0.06 | 0.00 | 0.04 | -0.19 | -0.43 | 0.05 | 0.13 |
| Maternal age |  | 0.01 | -0.01 | 0.03 | 0.27 | 0.00 | -0.01 | 0.00 | <0.0001 | -0.01 | -0.03 | 0.01 | 0.23 |
| Maternal height |  | 0.06 | 0.05 | 0.08 | <0.0001 | 0.00 | 0.00 | 0.00 | 0.02 | 0.02 | 0.01 | 0.04 | <0.0001 |
| Gravidity (ref: 1) | 2 | 0.07 | -0.21 | 0.35 | 0.65 | 0.08 | 0.05 | 0.11 | <0.0001 | 0.06 | -0.16 | 0.28 | 0.61 |
|  | >2 | 0.33 | 0.04 | 0.61 | 0.02 | 0.10 | 0.07 | 0.13 | <0.0001 | 0.11 | -0.12 | 0.33 | 0.35 |
| Sex (ref: male) | female | -0.62 | -0.83 | -0.42 | <0.0001 | 0.01 | -0.01 | 0.03 | 0.46 | -0.07 | -0.23 | 0.09 | 0.39 |
| Season (ref: spring) | summer | 0.09 | -0.21 | 0.39 | 0.55 | -0.01 | -0.04 | 0.02 | 0.56 | 0.04 | -0.19 | 0.28 | 0.72 |
|  | autumn | 0.21 | -0.08 | 0.51 | 0.16 | 0.01 | -0.02 | 0.04 | 0.60 | 0.13 | -0.11 | 0.36 | 0.28 |
|  | winter | 0.31 | 0.00 | 0.62 | 0.05 | 0.00 | -0.03 | 0.03 | 0.86 | 0.15 | -0.09 | 0.39 | 0.23 |
| Morphology (ref: none) | obese | 0.90 | 0.36 | 1.43 | <0.0001 | 0.01 | -0.05 | 0.06 | 0.75 | 0.57 | 0.16 | 0.99 | 0.01 |
|  | thin | -1.14 | -1.77 | -0.51 | <0.0001 | -0.09 | -0.15 | -0.02 | 0.01 | -0.95 | -1.44 | -0.46 | <0.0001 |
| Civil status (ref: married) | single or missing | -0.39 | -0.72 | -0.06 | 0.02 | -0.03 | -0.06 | 0.01 | 0.14 | -0.41 | -0.67 | -0.15 | <0.0001 |
| Living inside Lausanne (ref: yes) | no or unsure | 0.35 | 0.15 | 0.56 | <0.0001 | 0.01 | -0.01 | 0.03 | 0.37 | 0.28 | 0.11 | 0.44 | <0.0001 |
| *n* |  | 1,997 | | | | 1,997 | | | |  | 1,999 |  |  |

**Table S8: pregnancy outcomes depending on maternal ILI, multivariable GLMs for low birth weight, preterm birth (only using livebirths), full tables.** p: p-value.

|  |  | **Low birth weight (<2,500g)** | | | | **Preterm birth (<37 weeks)** | | | |
| --- | --- | --- | --- | --- | --- | --- | --- | --- | --- |
|  |  |  | 95% CI | |  |  | 95% CI | |  |
| Parameters | category | OR | lci | uci | pvalue | OR | lci | uci | pvalue |
| Influenza during pregnancy (ref: no) | yes | 2.06 | 1.33 | 3.20 | <0.0001 | 1.55 | 0.97 | 2.46 | 0.07 |
| Maternal age |  | 1.02 | 0.99 | 1.06 | 0.17 | 1.01 | 0.98 | 1.05 | 0.45 |
| Maternal height |  | 0.95 | 0.93 | 0.97 | <0.0001 | 0.95 | 0.93 | 0.98 | <0.0001 |
| Gravidity (ref: 1) | 2 | 0.80 | 0.49 | 1.30 | 0.37 | 0.93 | 0.58 | 1.50 | 0.76 |
|  | >2 | 0.81 | 0.50 | 1.29 | 0.37 | 0.85 | 0.53 | 1.38 | 0.52 |
| Sex (ref: male) | female | 0.80 | 0.57 | 1.14 | 0.21 | 1.07 | 0.75 | 1.51 | 0.71 |
| Season (ref: spring) | summer | 1.31 | 0.80 | 2.15 | 0.28 | 0.86 | 0.52 | 1.42 | 0.55 |
|  | autumn | 1.15 | 0.70 | 1.89 | 0.58 | 0.94 | 0.58 | 1.52 | 0.79 |
|  | winter | 0.79 | 0.45 | 1.37 | 0.40 | 0.77 | 0.45 | 1.30 | 0.32 |
| Morphology (ref: none) | obese | 0.36 | 0.09 | 1.52 | 0.17 | 0.19 | 0.03 | 1.37 | 0.10 |
|  | thin | 2.73 | 1.32 | 5.65 | 0.01 | 2.51 | 1.18 | 5.34 | 0.02 |
| Civil status (ref: married) | single or missing | 1.66 | 1.02 | 2.70 | 0.04 | 1.68 | 1.03 | 2.73 | 0.04 |
| Living inside Lausanne (ref: yes) | no or unsure | 0.73 | 0.52 | 1.03 | 0.08 | 0.62 | 0.44 | 0.88 | 0.01 |
| *n* |  | 1,999 | | | | 1,999 | | | |

**Table S9: Maternal characteristics depending on trimester of exposure to maternal ILI.** p: p-value. *^1^*Mean (SD); n (%). *^2^*Wilcoxon rank sum test; Fisher's exact test. note: the p-value compares exposure in each trimester vs. not exposed.

| **ILI** | **not exposed**, *n* = 1,895*^1^* | **first**, *n* = 75*^1^* | **p-value***^2^* | | **second**, *n* = 95*^1^* | **p-value***^2^* | **third,** *n* = 98*^1^* | **p-value***^2^* | **unknown**, *n* = 14*^1^* | **p-value***^2^* |
| --- | --- | --- | --- | --- | --- | --- | --- | --- | --- | --- |
| maternal age (years) | 28.5 (6.2) | 29.0 (6.7) | 0.7 | | 28.2 (5.9) | 0.7 | 28.3 (5.9) | 0.9 | 31.0 (6.0) | 0.11 |
| missing | 4 | 0 |  | | 0 |  | 0 |  | 0 |  |
| height (cm) | 156.4 (6.5) | 155.8 (6.4) | 0.2 | | 156.8 (6.5) | 0.9 | 156.0 (7.3) | 0.4 | 154.5 (7.8) | 0.5 |
| missing | 90 | 3 |  | | 3 |  | 5 |  | 0 |  |
| civil status |  |  | 0.3 | |  | 0.5 |  | 0.4 |  | 0.7 |
| married | 1,668 (88%) | 63 (84%) |  | | 86 (91%) |  | 89 (91%) |  | 12 (86%) |  |
| single or missing | 227 (12%) | 12 (16%) |  | | 9 (9%) |  | 9 (9%) |  | 2 (14%) |  |
| gravidity |  |  | 0.5 | |  | 0.6 |  | 0.2 |  | 0.3 |
| 1 | 758 (40%) | 35 (47%) |  | | 35 (37%) |  | 31 (32%) |  | 3 (21%) |  |
| 2 | 413 (22%) | 16 (21%) |  | | 25 (26%) |  | 26 (27%) |  | 3 (21%) |  |
| >2 | 724 (38%) | 24 (32%) |  | | 35 (37%) |  | 41 (42%) |  | 8 (57%) |  |
| living in Lausanne | 851 (45%) | 34 (45%) | >0.9 | | 45 (47%) | 0.6 | 47 (48%) | 0.6 | 4 (29%) | 0.2 |
| morphology |  |  | 0.7 | |  | 0.2 |  | 0.5 |  | >0.9 |
| neither | 1,770 (93%) | 69 (92%) |  | | 86 (91%) |  | 92 (94%) |  | 14 (100%) |  |
| obese | 72 (4%) | 4 (5%) |  | | 7 (7%) |  | 2 (2%) |  | 0 (0%) |  |
| thin | 53 (3%) | 2 (3%) |  | | 2 (2%) |  | 4 (4%) |  | 0 (0%) |  |
| goitre | 106 (6%) | 2 (3%) | 0.4 | | 6 (6%) | 0.8 | 6 (6%) | 0.8 | 2 (14%) | 0.2 |
| rickets | 76 (4%) | 4 (5%) | 0.5 | | 3 (3%) | >0.9 | 3 (3%) | >0.9 | 1 (7%) | 0.4 |
| syphilis | 20 (1%) | 3 (4%) | 0.054 | | 1 (1%) | >0.9 | 0 (0%) | 0.6 | 0 (0%) | >0.9 |
| HISCO class |  |  | 0.6 | |  | 0.6 |  | >0.9 |  | 0.9 |
| 2 | 1,627 (86%) | 63 (84%) |  | | 86 (91%) |  | 86 (88%) |  | 14 (100%) |  |
| 1 | 112 (6%) | 6 (8%) |  | | 3 (3%) |  | 6 (6%) |  | 0 (0%) |  |
| 3 | 34 (2%) | 2 (3%) |  | | 2 (2%) |  | 1 (1%) |  | 0 (0%) |  |
| missing | 122 (6%) | 4 (5%) |  | | 4 (4%) |  | 5 (5%) |  | 0 (0%) |  |
| season |  |  | <0.001 | |  | <0.001 |  | <0.001 |  | 0.002 |
| spring | 344 (18%) | 29 (39%) |  | | 43 (45%) |  | 23 (23%) |  | 7 (50%) |  |
| summer | 502 (26%) | 34 (45%) |  | | 18 (19%) |  | 10 (10%) |  | 5 (36%) |  |
| autumn | 628 (33%) | 7 (9.3%) |  | | 7 (7.4%) |  | 24 (24%) |  | 0 (0%) |  |
| winter | 421 (22%) | 5 (6.7%) |  | | 27 (28%) |  | 41 (42%) |  | 2 (14%) |  |
|  |  |  | | *^1^*Mean (SD); n (%) | | | | | |  |

**Table S10: pregnancy outcomes depending on trimester of exposure to maternal ILI among males (*n*=1,122).** p: p-value. *^1^*Mean (SD); n (%). *^2^*Wilcoxon rank sum test; Fisher's exact test. note: the p-value compares exposure in each trimester vs. not exposed

| **Trimester of exposure** | **not exposed**,  *n* = 992*^1^* | **first**,  *n*= 31*^1^* | **p^2^** | **second**,  *n* = 48*^1^* | **p^2^** | **third**,  *n* = 46*^1^* | **p^2^** | **unknown**,  *n* = 5*^1^* |
| --- | --- | --- | --- | --- | --- | --- | --- | --- |
| birth weight (g) | 3,263.6 (584.8) | **3,038.4 (615.6)** | **0.036** | 3,310.6 (569.2) | 0.4 | **2,918.0 (694.5)** | **0.001** | 3,126.0 (715.9) |
| head circumference (cm) | 34.7 (1.8) | 34.1 (2.2) | 0.11 | 34.5 (1.9) | 0.5 | **33.9 (2.4)** | **0.018** | 33.6 (1.9) |
| placenta weight (g) | 595.7 (124.0) | **545.6 (94.0)** | **0.012** | 598.5 (108.3) | 0.6 | **508.2 (112.1)** | **<0.001** | 512.0 (111.2) |
| birth length (cm) | 49.5 (2.7) | **48.2 (3.5)** | **0.014** | 49.8 (2.7) | 0.3 | **48.2 (4.0)** | **0.017** | 48.6 (2.7) |
| ponderal index | 2.7 (0.3) | 2.7 (0.2) | >0.9 | 2.6 (0.2) | 0.5 | **2.6 (0.3)** | **0.002** | 2.7 (0.2) |
| gestational age (weeks) | 39.3 (2.1) | 39.0 (2.7) | >0.9 | 39.5 (2.1) | 0.2 | **38.1 (3.2)** | **0.013** | 39.3 (2.7) |
| low birth weight (<2'500g) | 85 (9%) | 5 (16%) | 0.2 | 3 (6%) | 0.8 | **12 (26%)** | **<0.001** | 1 (20%) |
| preterm birth (<37 weeks) | 78 (8%) | 3 (10%) | 0.7 | 4 (8%) | 0.8 | **10 (22%)** | **0.004** | 1 (20%) |
| stillbirth | 45 (5%) | 0 (0%) | 0.4 | 2 (4%) | >0.9 | **6 (13%)** | **0.022** | 0 (0%) |
| neonatal mortality d1-5 | 17 (2%) | 0 (0%) | >0.9 | 0 (0%) | >0.9 | 2 (4%) | 0.2 | 0 (0%) |

**Table S11: pregnancy outcomes depending on trimester of exposure to maternal ILI among females.** p: p-value. *^1^*Mean (SD); n (%). *^2^*Wilcoxon rank sum test; Fisher's exact test. note: the p-value compares exposure in each trimester vs. not exposed.

| **ILI exposure** | **not exposed**,  *n* = 898*^1^* | **first**,  *n*= 44*^1^* | **p^2^** | **second**,  *n* = 47*^1^* | **p^2^** | **third**,  *n* = 52*^1^* | **p^2^** | **unknown**,  *n* = 9*^1^* |
| --- | --- | --- | --- | --- | --- | --- | --- | --- |
| birth weight (g) | 3,142.2 (520.0) | 3,104.8 (552.5) | 0.6 | 3,087.4 (485.5) | 0.2 | 3,014.6 (569.9) | 0.10 | 3,107.8 (468.6) |
| head circumference (cm) | 34.1 (1.7) | 34.2 (1.6) | 0.7 | 34.2 (2.1) | 0.8 | 33.7 (1.6) | 0.094 | 33.9 (1.5) |
| placenta weight (g) | 576.9 (112.8) | 592.8 (113.7) | 0.5 | **532.1 (93.6)** | **0.003** | **533.3 (91.4)** | **0.003** | 526.7 (117.5) |
| birth length (cm) | 48.8 (2.5) | 48.7 (1.8) | 0.5 | 48.9 (2.1) | 0.6 | 48.3 (3.1) | 0.3 | 48.3 (3.2) |
| ponderal index | 2.7 (0.3) | 2.7 (0.3) | >0.9 | 2.6 (0.3) | 0.3 | 2.7 (0.2) | 0.3 | 2.8 (0.3) |
| gestational age (weeks) | 39.2 (2.0) | 39.6 (1.4) | 0.2 | 39.3 (1.4) | 0.6 | **38.6 (2.6)** | **0.009** | 38.9 (2.6) |
| low birth weight (<2'500g) | 70 (8%) | 6 (14%) | 0.2 | 4 (9%) | 0.8 | 6 (12%) | 0.3 | 1 (11%) |
| preterm birth (<37 weeks) | 73 (8%) | 2 (5%) | 0.6 | 3 (6%) | >0.9 | 8 (15%) | 0.075 | 2 (22%) |
| stillbirth | 33 (4%) | 1 (2%) | >0.9 | 0 (0%) | 0.4 | 3 (6%) | 0.4 | 0 (0%) |
| neonatal mortality d1-5 | 20 (2%) | 0 (0%) | 0.6 | 2 (4%) | 0.3 | 2 (4%) | 0.3 | 0 (0%) |

**Table S12: pregnancy outcomes depending on trimester of exposure to maternal ILI and infant sex, univariable GLMs.** All models are fitted using only livebirths, except for stillbirth outcome. Note: when the timing of ILI exposure was unknown (*n*=14), cases were not considered. For each outcome, only one GLM was fitted, and the exposure variable was a categorical variable (no ILI (reference category), ILI in the first, second or third trimester). p: p-value.

|  |  | **Univariable (males and females)** | | | | | **Univariable Males** | | | | | **Univariable Females** | | | | |
| --- | --- | --- | --- | --- | --- | --- | --- | --- | --- | --- | --- | --- | --- | --- | --- | --- |
| **Continuous variables** |  |  | 95% CI | |  |  |  | 95% CI | |  | |  | 95% CI | |  | |
|  | trimester | beta | lci | uci | p-value | *n* | beta | lci | uci | p-value | *n* | beta | lci | uci | p-value | *n* |
| Birth weight (g) | first | **-134.50** | **-256.83** | **-12.18** | **0.03** | 2,071 | **-254.09** | **-450.37** | **-57.82** | **0.01** | 1,064 | -25.97 | -177.35 | 125.41 | 0.74 | 1,004 |
|  | second | -14.60 | -124.26 | 95.06 | 0.79 |  | 58.39 | -103.97 | 220.74 | 0.48 |  | -79.22 | -224.34 | 65.89 | 0.28 |  |
|  | third | **-218.41** | **-330.39** | **-106.42** | **<0.0001** |  | **-268.98** | **-442.56** | **-95.40** | **<0.0001** |  | **-160.55** | **-302.83** | **-18.27** | **0.03** |  |
| Head circumference (cm) | first | -0.22 | -0.60 | 0.16 | 0.25 | 2,063 | **-0.68** | **-1.27** | **-0.09** | **0.02** | 1.061 | 0.23 | -0.25 | 0.71 | 0.35 | 999 |
|  | second | 0.04 | -0.31 | 0.38 | 0.84 |  | -0.03 | -0.52 | 0.46 | 0.91 |  | 0.14 | -0.32 | 0.60 | 0.55 |  |
|  | third | **-0.56** | **-0.91** | **-0.21** | **<0.0001** |  | **-0.59** | **-1.12** | **-0.06** | **0.03** |  | -0.44 | -0.89 | 0.02 | 0.06 |  |
| Placenta weight (g) | first | -12.60 | -39.10 | 13.89 | 0.35 | 2,045 | **-53.28** | **-95.40** | **-11.17** | **0.01** | 1,048 | 20.42 | -12.77 | 53.61 | 0.23 | 994 |
|  | second | -23.35 | -47.22 | 0.53 | 0.06 |  | 1.07 | -34.14 | 36.28 | 0.95 |  | -45.46 | -77.27 | -13.64 | 0.01 |  |
|  | third | **-65.59** | **-89.85** | **-41.34** | **<0.0001** |  | **-84.93** | **-122.18** | **-47.68** | **<0.0001** |  | **-47.09** | **-78.29** | **-15.90** | **<0.0001** |  |
| Birth length (cm) | first | **-0.72** | **-1.28** | **-0.15** | **0.01** | 2,067 | **-1.37** | **-2.25** | **-0.49** | **<0.0001** | 1,063 | -0.13 | -0.85 | 0.60 | 0.73 | 1,001 |
|  | second | 0.23 | -0.28 | 0.73 | 0.37 |  | 0.49 | -0.24 | 1.22 | 0.19 |  | 0.01 | -0.68 | 0.69 | 0.99 |  |
|  | third | **-0.81** | **-1.32** | **-0.29** | **<0.0001** |  | **-0.81** | **-1.58** | **-0.03** | **0.04** |  | **-0.72** | **-1.40** | **-0.05** | **0.04** |  |
| Ponderal index | first | 0.00 | -0.06 | 0.05 | 0.90 | 2,067 | -0.01 | -0.09 | 0.08 | 0.89 | 1,063 | 0.00 | -0.08 | 0.07 | 0.96 | 1,001 |
|  | second | -0.04 | -0.10 | 0.01 | 0.08 |  | -0.03 | -0.10 | 0.05 | 0.49 |  | -0.06 | -0.13 | 0.01 | 0.08 |  |
|  | third | **-0.07** | **-0.12** | **-0.01** | **0.01** |  | **-0.11** | **-0.18** | **-0.03** | **0.01** |  | -0.03 | -0.10 | 0.04 | 0.35 |  |
| Gestational age (weeks) | first | -0.05 | -0.48 | 0.38 | 0.81 | 2,071 | -0.44 | -1.11 | 0.23 | 0.20 | 1,064 | 0.23 | -0.32 | 0.79 | 0.41 | 1,004 |
|  | second | 0.16 | -0.23 | 0.54 | 0.43 |  | 0.31 | -0.24 | 0.86 | 0.27 |  | 0.01 | -0.53 | 0.54 | 0.98 |  |
|  | third | **-0.76** | **-1.16** | **-0.37** | **<0.0001** |  | **-0.69** | **-1.28** | **-0.10** | **0.02** |  | **-0.82** | **-1.34** | **-0.29** | **<0.0001** |  |
| **Binary variables** |  |  | 95% CI | |  |  |  | 95% CI | |  | |  | 95% CI | |  | |
|  |  | OR | lci | uci | p-value | *n* | OR | lci | uci | p-value | *n* | OR | lci | uci | p-value | *n* |
| Low birth weight (<2’500g) | first | **2.11** | **1.06** | **4.22** | **0.03** | 2,071 | 2.45 | 0.91 | 6.57 | 0.08 | 1,064 | 1.90 | 0.72 | 5.02 | 0.19 | 1,004 |
|  | second | 0.93 | 0.40 | 2.18 | 0.87 |  | 0.58 | 0.14 | 2.44 | 0.46 |  | 1.34 | 0.47 | 3.88 | 0.58 |  |
|  | third | **2.52** | **1.39** | **4.59** | **<0.0001** |  | **3.18** | **1.41** | **7.17** | **0.01** |  | **2.02** | **0.82** | **4.94** | **0.13** |  |
| Preterm birth (<37 weeks) | first | 1.01 | 0.40 | 2.54 | 0.99 | 2,071 | 1.50 | 0.44 | 5.08 | 0.51 | 1,064 | 0.67 | 0.16 | 2.82 | 0.58 | 1,004 |
|  | second | 0.96 | 0.41 | 2.23 | 0.92 |  | 0.98 | 0.30 | 3.24 | 0.97 |  | 0.93 | 0.28 | 3.09 | 0.91 |  |
|  | third | **2.59** | **1.42** | **4.72** | **<0.0001** |  | **2.48** | **1.00** | **6.12** | **0.05** |  | **2.67** | **1.19** | **5.95** | **0.02** |  |
| Stillbirth | first | 0.31 | 0.04 | 2.23 | 0.24 | 2,163 | 0.00 | 0.00 | Inf | 0.98 | 1,117 | 0.61 | 0.08 | 4.56 | 0.63 |  |
|  | second | 0.49 | 0.12 | 2.02 | 0.32 |  | 0.91 | 0.22 | 3.89 | 0.90 |  | 0.00 | 0.00 | Inf | 0.99 | 1,041 |
|  | third | **2.29** | **1.12** | **4.72** | **0.02** |  | **3.16** | **1.27** | **7.83** | **0.01** |  | 1.60 | 0.48 | 5.42 | 0.45 |  |
| Neonatal mortality day 1-5 | first | 0.00 | 0.00 | Inf | 0.98 | 2,070 | 0.00 | 0.00 | Inf | 0.99 | 1,064 | 0.00 | 0.00 | Inf | 0.99 | 1,003 |
|  | second | 1.06 | 0.25 | 4.45 | 0.94 |  | 0.00 | 0.00 | Inf | 0.99 |  | 1.88 | 0.43 | 8.28 | 0.41 |  |
|  | third | 2.29 | 0.80 | 6.57 | 0.12 |  | 2.88 | 0.64 | 12.91 | 0.17 |  | 1.84 | 0.42 | 8.10 | 0.42 |  |

**Table S13: pregnancy outcomes depending on trimester of exposure to maternal ILI, multivariable GLMs.** All models are fitted using only livebirths. Note: when the timing of ILI exposure was unknown (*n*=14), cases were not considered. For each outcome, only one GLM was fitted, and the exposure variable was a categorical variable (no ILI (reference category), ILI in the first, second or third trimester). Models were not stratified by sex due to small sample size. p: p-value.

|  |  | **Multivariable (males and females)** | | | | |
| --- | --- | --- | --- | --- | --- | --- |
| **Continuous variables** |  |  | 95% CI | |  |  |
|  | trimester | beta | lci | uci | p-value | *n* |
| Birth weight (g) | first | -61.09 | -180.81 | 58.62 | 0.32 | 1,985 |
|  | second | -1.95 | -108.97 | 105.07 | 0.97 |  |
|  | third | **-214.45** | **-323.75** | **-105.16** | **<0.0001** |  |
| Head circumference (cm) | first | 0.02 | -0.36 | 0.40 | 0.91 | 1,978 |
|  | second | 0.10 | -0.24 | 0.44 | 0.57 |  |
|  | third | **-0.49** | **-0.84** | **-0.14** | **0.01** |  |
| Placenta weight (g) | first | -2.39 | -28.98 | 24.19 | 0.86 | 1,963 |
|  | second | -20.62 | -44.50 | 3.27 | 0.09 |  |
|  | third | **-59.60** | **-83.87** | **-35.33** | **<0.0001** |  |
| Birth length (cm) | first | -0.44 | -1.00 | 0.12 | 0.12 | 1,983 |
|  | second | 0.27 | -0.23 | 0.77 | 0.28 |  |
|  | third | **-0.78** | **-1.28** | **-0.27** | **<0.0001** |  |
| Ponderal index | first | 0.02 | -0.04 | 0.07 | 0.60 | 1,983 |
|  | second | -0.04 | -0.10 | 0.01 | 0.09 |  |
|  | third | **-0.07** | **-0.12** | **-0.02** | **0.01** |  |
| Gestational age (weeks) | first | 0.09 | -0.34 | 0.53 | 0.68 | 1,985 |
|  | second | 0.18 | -0.20 | 0.57 | 0.35 |  |
|  | third | **-0.79** | **-1.19** | **-0.39** | **<0.0001** |  |
| **Binary variables** |  |  | 95% CI | |  |  |
|  |  | OR | lci | uci | p-value | *n* |
| Low birth weight (<2’500g) | first | 1.86 | 0.88 | 3.93 | 0.10 | 1,985 |
|  | second | 1.18 | 0.49 | 2.81 | 0.71 |  |
|  | third | **3.27** | **1.74** | **6.16** | **<0.0001** |  |
| Preterm birth (<37 weeks) | first | 0.74 | 0.26 | 2.12 | 0.58 | 1,985 |
|  | second | 0.85 | 0.33 | 2.17 | 0.73 |  |
|  | third | **2.87** | **1.53** | **5.39** | **<0.0001** |  |

**Table S14: Maternal characteristics depending on the severity of ILI symptoms.** p: p-value. *^1^*Mean (SD); n (%). *^2^*Wilcoxon rank sum test; Fisher's exact test. note: the p-value compares symptoms severity vs. not exposed.

| **ILI exposure** | **not exposed**, *n* = 1,895*^1^* | **mild symptoms**, *n* = 137*^1^* | **p***^2^* | **severe symptoms**, *n* = 100*^1^* | **p***^2^* | **symptoms unknown**, *n* = 45*^1^* | **p***^2^* |
| --- | --- | --- | --- | --- | --- | --- | --- |
| maternal age (years) | 28.5 (6.2) | 29.4 (6.3) | 0.11 | 27.7 (5.6) | 0.3 | 27.9 (6.5) | 0.5 |
| missing | 4 | 0 |  | 0 |  | 0 |  |
| height (cm) | 156.4 (6.5) | 156.1 (6.9) | 0.3 | 155.5 (6.9) | 0.2 | 157.8 (6.4) | 0.4 |
| missing | 90 | 4 |  | 3 |  | 4 |  |
| civil status |  |  | 0.5 |  | 0.4 |  | 0.5 |
| married | 1,668 (88%) | 118 (86%) |  | 91 (91%) |  | 41 (91%) |  |
| single or missing | 227 (12%) | 19 (14%) |  | 9 (9%) |  | 4 (9%) |  |
| gravidity |  |  | >0.9 |  | **0.018** |  | 0.3 |
| 1 | 758 (40%) | 53 (39%) |  | **28 (28%)** |  | 23 (51%) |  |
| 2 | 413 (22%) | 31 (23%) |  | **32 (32%)** |  | 7 (16%) |  |
| >2 | 724 (38%) | 53 (39%) |  | **40 (40%)** |  | 15 (33%) |  |
| living in Lausanne | 851 (45%) | 62 (45%) | >0.9 | 42 (42%) | 0.6 | 26 (58%) | 0.086 |
| morphology |  |  | >0.9 |  | 0.4 |  | 0.9 |
| neither | 1,770 (93%) | 128 (93%) |  | 91 (91%) |  | 42 (93%) |  |
| obese | 72 (4%) | 5 (4%) |  | 6 (6%) |  | 2 (4%) |  |
| thin | 53 (3%) | 4 (3%) |  | 3 (3%) |  | 1 (2%) |  |
| goitre | 106 (6%) | 9 (7%) | 0.6 | 6 (6%) | 0.9 | 1 (2%) | 0.5 |
| rickets | 76 (4%) | 4 (3%) | 0.5 | 6 (6%) | 0.3 | 1 (2%) | >0.9 |
| syphilis | 20 (1%) | 3 (2%) | 0.2 | 0 (0%) | 0.6 | 1 (2%) | 0.4 |
| HISCO class |  |  | 0.7 |  | 0.8 |  | 0.3 |
| 2 | 1,627 (86%) | 123 (90%) |  | 90 (90%) |  | 36 (80%) |  |
| 1 | 112 (6%) | 6 (4%) |  | 5 (5%) |  | 4 (9%) |  |
| 3 | 34 (2%) | 2 (1%) |  | 1 (1%) |  | 2 (4%) |  |
| missing | 122 (6%) | 6 (4%) |  | 4 (4%) |  | 3 (7%) |  |
| season |  |  | <0.001 |  | <0.001 |  | 0.043 |
| spring | 344 (18%) | 64 (47%) |  | 31 (31%) |  | 7 (16%) |  |
| summer | 502 (26%) | 29 (21%) |  | 18 (18%) |  | 20 (44%) |  |
| autumn | 628 (33%) | 11 (8.0%) |  | 14 (14%) |  | 13 (29%) |  |
| winter | 421 (22%) | 33 (24%) |  | 37 (37%) |  | 5 (11%) |  |

**Table S15: maternal ILI depending on trimester of illness and symptoms severity**

|  | trimester of ILI | | | |
| --- | --- | --- | --- | --- |
| symptoms severity | first | second | third | unknown |
| mild | 49 (65,3%) | 50 (52,6%) | 32 (32,7%) | 6 (42,9%) |
| severe | 11 (14,7%) | 35 (36,8%) | 48 (49,0%) | 6 (42,9%) |
| unknown | 15 (20,0%) | 10 (10,5%) | 18 (18,4%) | 2 (14,3%) |
| total | 75 (100%) | 95 (100%) | 98 (100%) | 14 (100%) |

**Table S16: pregnancy outcomes depending on the severity of ILI symptoms among males (*n*=1,112).** p: p-value. *^1^*Mean (SD); n (%). *^2^*Wilcoxon rank sum test; Fisher's exact test. note: the p-value compares exposure to ILI with mild symptoms vs. not exposed.

| **ILI exposure** | **not infected** | **mild** | | **severe** | | **missing** |
| --- | --- | --- | --- | --- | --- | --- |
| **Variable** | *n* = 992*^1^* | *n* = 59*^1^* | **p^1^** | *n* = 53*^1^* | **p^1^** | *n* = 18*^1^* |
| birth weight (g) | 3,263.6 (584.8) | 3,123.1 (603.9) | 0.10 | 3,115.3 (687.7) | 0.2 | 2,977.2 (688.8) |
| head circumference (cm) | 34.7 (1.8) | 34.2 (2.1) | 0.093 | 34.3 (2.1) | 0.2 | 33.7 (2.4) |
| placenta weight (g) | 595.7 (124.0) | **547.7 (117.7)** | **0.006** | 556.0 (110.0) | 0.065 | 544.4 (107.1) |
| birth length (cm) | 49.5 (2.7) | 49.0 (3.3) | 0.3 | 48.9 (3.6) | 0.4 | 48.0 (3.4) |
| ponderal index | 2.7 (0.3) | 2.6 (0.2) | 0.081 | 2.6 (0.3) | 0.2 | 2.6 (0.3) |
| gestational age (weeks) | 39.3 (2.1) | 39.0 (2.3) | >0.9 | 39.3 (2.1) | 38.8 (3.1) | 0.8 |
| low birth weight (<2'500g) | 85 (9%) | 7 (12%) | 0.4 | **10 (19%)** | **0.023** | 4 (22%) |
| preterm birth (<37 weeks) | 78 (8%) | 8 (14%) | 0.14 | 8 (15%) | 0.071 | 2 (11%) |
| stillbirth | 45 (5%) | 2 (3%) | >0.9 | **6 (11%)** | **0.039** | 0 (0%) |
| neonatal mortality d1-5 | 17 (2%) | 1 (2%) | >0.9 | 1 (2%) | 0.6 | 0 (0%) |

**Table S17: pregnancy outcomes depending on the severity of ILI symptoms among females (*n*=1,050).** p: p-value. *^1^*Mean (SD); n (%). *^2^*Wilcoxon rank sum test; Fisher's exact test. note: the p-value compares exposure to ILI with severe symptoms vs. not exposed.

| **ILI exposure** | **not infected** | **mild** | | **severe** | | **missing** |
| --- | --- | --- | --- | --- | --- | --- |
| **Variable** | *n* = 898*^1^* | *n* = 78*^1^* | **p^1^** | *n* = 47*^1^* | **p^1^** | *n* = 27*^1^* |
| birth weight (g) | 3,142.2 (520.0) | 3,069.2 (464.3) | 0.079 | 3,059.4 (606.6) | 0.4 | 2,977.2 (688.8) |
| head circumference (cm) | 34.1 (1.7) | 34.0 (1.4) | 0.3 | 34.2 (2.4) | >0.9 | 33.7 (2.4) |
| placenta weight (g) | 576.9 (112.8) | 552.9 (108.1) | 0.052 | 548.6 (95.2) | 0.059 | 544.4 (107.1) |
| birth length (cm) | 48.8 (2.5) | 48.7 (2.2) | 0.3 | 48.6 (2.8) | 0.8 | 48.0 (3.4) |
| ponderal index | 2.7 (0.3) | 2.7 (0.3) | 0.5 | 2.6 (0.2) | 0.2 | 2.6 (0.3) |
| gestational age (weeks) | 39.2 (2.0) | 39.3 (1.7) | 0.8 | **38.8 (2.3)** | **0.040** | 39.1 (2.3) |
| low birth weight (<2'500g) | 70 (8%) | 6 (8%) | >0.9 | 7 (15%) | 0.10 | 4 (22%) |
| preterm birth (<37 weeks) | 73 (8%) | 7 (9%) | 0.8 | 6 (13%) | 0.3 | 2 (11%) |
| stillbirth | 33 (4%) | 3 (4%) | 0.8 | 1 (2%) | >0.9 | 0 (0%) |
| neonatal mortality d1-5 | 20 (2%) | 0 (0%) | 0.4 | 3 (7%) | 0.10 | 0 (0%) |

**Table S18: pregnancy outcomes depending on the severity of ILI symptoms and infant sex, univariable GLMs.** All models are fitted using only livebirths, except for stillbirth outcome. Note: when the severity of ILI was unknown (*n*=45), cases were not considered. For each outcome, only one GLM was fitted, and the exposure variable was a categorical variable (no ILI (reference category), ILI with mild symptoms, ILI with severe symptoms). p: p-value.

|  |  | **Univariable (males and females)** | | | | | **Univariable Males** | | | | | **Univariable Females** | | | | |
| --- | --- | --- | --- | --- | --- | --- | --- | --- | --- | --- | --- | --- | --- | --- | --- | --- |
| **Continuous variables** |  |  | 95% CI | |  |  |  | 95% CI | |  | |  | 95% CI | |  | |
|  | severity | beta | lci | uci | p | *n* | beta | lci | uci | p | *n* | beta | lci | uci | p | *n* |
| Birth weight (g) | mild | **-121.58** | **-214.28** | **-28.89** | **0.01** | 2,040 | -143.53 | -290.11 | 3.04 | 0.06 | 1,051 | -84.94 | -200.92 | 31.04 | 0.15 | 986 |
|  | severe | -83.31 | -192.63 | 26.01 | 0.14 |  | -62.27 | -222.88 | 98.34 | 0.45 |  | -100.58 | -246.37 | 45.21 | 0.18 |  |
| Head circumference (cm) | mild | **-0.30** | **-0.59** | **-0.01** | **0.05** | 2,032 | -0.40 | -0.83 | 0.04 | 0.08 | 1,048 | -0.11 | -0.48 | 0.26 | 0.57 | 981 |
|  | severe | -0.07 | -0.42 | 0.27 | 0.68 |  | -0.20 | -0.68 | 0.28 | 0.42 |  | 0.08 | -0.39 | 0.56 | 0.73 |  |
| Placenta weight (g) | mild | **-37.61** | **-57.90** | **-17.31** | **<0.0001** | 2,014 | **-51.87** | **-83.81** | **-19.94** | **<0.0001** | 1,035 | -23.52 | -49.27 | 2.22 | 0.07 | 976 |
|  | severe | **-31.32** | **-55.16** | **-7.47** | **0.01** |  | -32.97 | -67.67 | 1.73 | 0.06 |  | -29.02 | -61.38 | 3.34 | 0.08 |  |
| Birth length (cm) | mild | -0.38 | -0.80 | 0.05 | 0.09 | 2,036 | -0.41 | -1.06 | 0.24 | 0.22 | 1,050 | -0.25 | -0.81 | 0.30 | 0.37 | 983 |
|  | severe | -0.18 | -0.68 | 0.32 | 0.48 |  | -0.13 | -0.84 | 0.59 | 0.73 |  | -0.21 | -0.91 | 0.48 | 0.55 |  |
| Ponderal index | mild | -0.03 | -0.08 | 0.01 | 0.12 | 2,036 | -0.05 | -0.12 | 0.01 | 0.12 | 1,050 | -0.02 | -0.08 | 0.04 | 0.48 | 983 |
|  | severe | **-0.05** | **-0.10** | **0.00** | **0.04** |  | -0.04 | -0.11 | 0.03 | 0.29 |  | -0.07 | -0.14 | 0.00 | 0.06 |  |
| Gestational age (weeks) | mild | -0.12 | -0.45 | 0.20 | 0.46 | 2,040 | -0.21 | -0.71 | 0.28 | 0.40 | 1,051 | -0.04 | -0.47 | 0.39 | 0.84 | 986 |
|  | severe | -0.26 | -0.65 | 0.12 | 0.18 |  | -0.02 | -0.56 | 0.52 | 0.94 |  | -0.51 | -1.05 | 0.03 | 0.07 |  |
| **Binary variables** |  |  | 95% CI | |  |  |  | 95% CI | |  | |  | 95% CI | |  | |
|  |  | OR | lci | uci | p-value | *n* | OR | lci | uci | p-value | *n* | OR | lci | uci | p-value | *n* |
| Low birth weight (<2’500g) | mild | 1.23 | 0.65 | 2.34 | 0.53 | 2,040 | 1.50 | 0.62 | 3.61 | 0.37 | 1,051 | 1.03 | 0.40 | 2.66 | 0.95 | 986 |
|  | severe | **2.20** | **1.19** | **4.06** | **0.01** |  | 1.86 | 0.76 | 4.54 | 0.17 |  | **2.59** | **1.11** | **6.06** | **0.03** |  |
| Preterm birth (<37 weeks) | mild | 1.65 | 0.92 | 2.95 | 0.09 | 2,040 | 1.96 | 0.86 | 4.51 | 0.11 | 1,051 | 1.41 | 0.62 | 3.20 | 0.42 | 986 |
|  | severe | 1.67 | 0.85 | 3.30 | 0.14 |  | 1.31 | 0.45 | 3.75 | 0.62 |  | 2.05 | 0.83 | 5.03 | 0.12 |  |
| Stillbirth | mild | 0.86 | 0.34 | 2.16 | 0.75 | 2,132 | 0.74 | 0.17 | 3.12 | 0.68 | 1,104 | 1.05 | 0.31 | 3.50 | 0.94 | 1,023 |
|  | severe | 1.71 | 0.77 | 3.80 | 0.19 |  | **2.69** | **1.09** | **6.61** | **0.03** |  | 0.57 | 0.08 | 4.26 | 0.58 |  |
| Neonatal mortality day 1-5 | mild | 0.37 | 0.05 | 2.69 | 0.32 | 2,039 | 0.98 | 0.13 | 7.47 | 0.98 | 1,051 | 0.00 | 0.00 | Inf | 0.99 | 985 |
|  | severe | 2.18 | 0.76 | 6.26 | 0.15 |  | 1.19 | 0.15 | 9.13 | 0.87 |  | 3.02 | 0.86 | 10.56 | 0.08 |  |

**Table S19: pregnancy outcomes depending on the severity of ILI symptoms and infant sex, multivariable GLMs.** All models are fitted using only livebirths. Note: when the severity of ILI was unknown (*n*=45), cases were not considered. For each outcome, only one GLM was fitted, and the exposure variable was a categorical variable (no ILI (reference category), ILI with mild symptoms, ILI with severe symptoms). Models were not stratified by sex due to small sample size. p: p-value.

|  |  | **Multivariable***^1^* | | | | |
| --- | --- | --- | --- | --- | --- | --- |
| **Continuous variables** |  |  | 95% CI | |  | |
|  | severity | beta | lci | uci | p | *n* |
| Birth weight (g) | mild | -64.52 | -155.36 | 26.32 | 0.16 | 1,958 |
|  | severity | -98.68 | -204.14 | 6.77 | 0.07 |  |
| Head circumference (cm) | mild | -0.08 | -0.37 | 0.21 | 0.58 | 1,951 |
|  | severity | -0.08 | -0.42 | 0.25 | 0.62 |  |
| Placenta weight (g) | mild | **-29.22** | **-49.66** | **-8.79** | **0.01** | 1,936 |
|  | severity | **-29.54** | **-53.19** | **-5.90** | **0.01** |  |
| Birth length (cm) | mild | -0.14 | -0.56 | 0.29 | 0.53 | 1,956 |
|  | severity | -0.21 | -0.70 | 0.29 | 0.41 |  |
| Ponderal index | mild | -0.02 | -0.07 | 0.02 | 0.27 | 1,956 |
|  | severity | **-0.07** | **-0.12** | **-0.01** | **0.01** |  |
| Gestational age (weeks) | mild | 0.01 | -0.32 | 0.34 | 0.95 | 1,958 |
|  | severity | -0.30 | -0.69 | 0.08 | 0.12 |  |
| **Binary variables** |  |  | 95% CI | |  | |
|  |  | OR | lci | uci | p | *n* |
| Low birth weight (<2’500g) | mild | 1.14 | 0.56 | 2.29 | 0.72 | 1,958 |
|  | severity | **2.75** | **1.44** | **5.23** | **<0.0001** |  |
| Preterm birth (<37 weeks) | mild | 1.31 | 0.68 | 2.52 | 0.42 | 1,958 |
|  | severity | 1.88 | 0.93 | 3.81 | 0.08 |  |
| Stillbirth | mild | - | | | | |
|  | severity |  |  |  |  |  |
| Neonatal mortality day 1-5 | mild | - | | | | |
|  | severity |  |  |  |  |  |

**MISCARRIAGES**

**Figure S6: yearly and quarterly miscarriage and birth counts between 1909 and 1921, and percentage of miscarriages relative to births.**

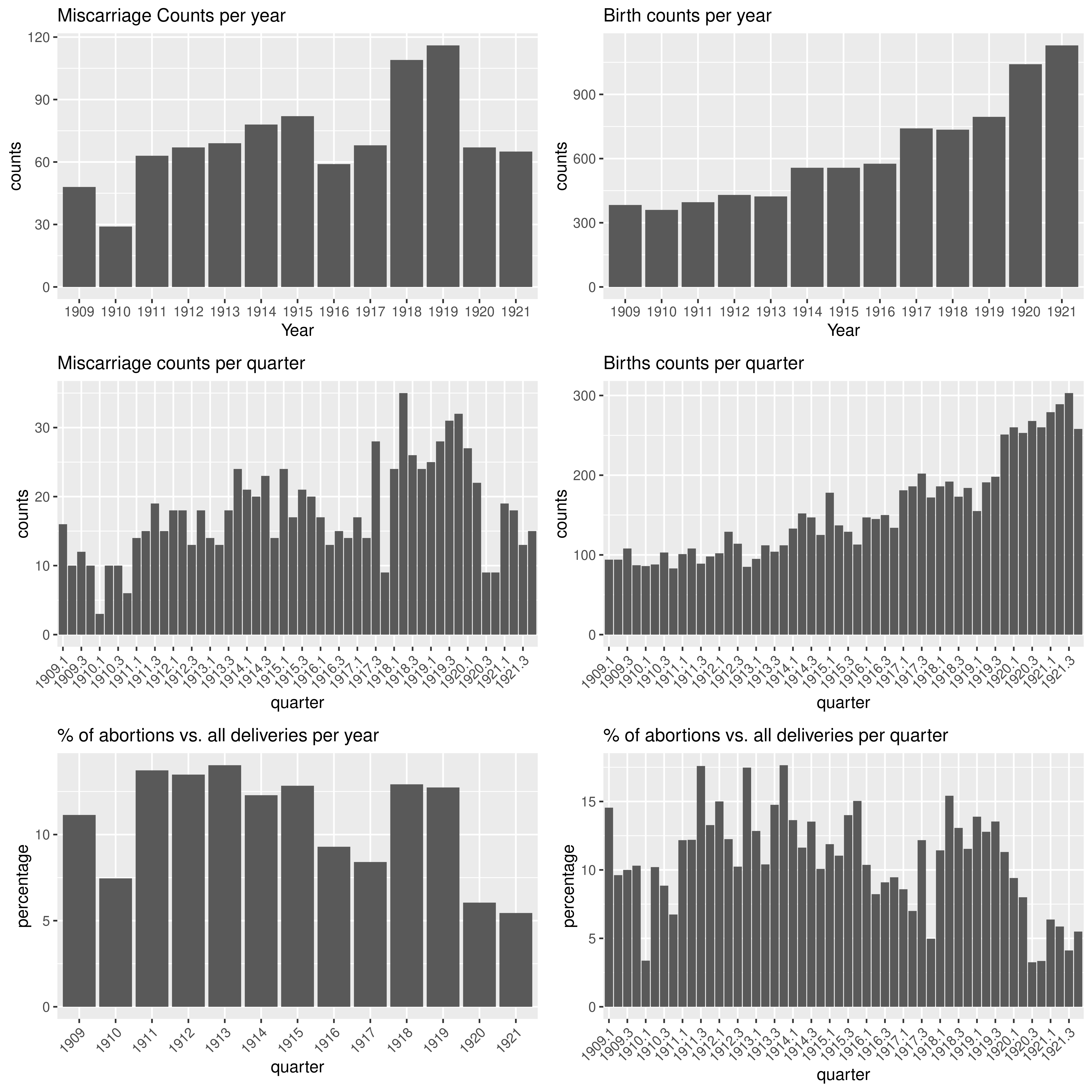

**Table S20: maternal characteristics of those who had a miscarriage, by year (1909-1920).**

| **Variable** | **1909**, N = 48*^1^* | **1910**, N = 29*^1^* | **1911**, N = 63*^1^* | **1912**, N = 67*^1^* | **1913**, N = 69*^1^* | **1914**, N = 78*^1^* | **1915**, N = 82*^1^* | **1916**, N = 59*^1^* | **1917**, N = 68*^1^* | **1918**, N = 109*^1^* | **1919**, N = 116*^1^* | **1920**, N = 67*^1^* | **1921**, N = 65*^1^* |
| --- | --- | --- | --- | --- | --- | --- | --- | --- | --- | --- | --- | --- | --- |
| maternal age (years) | 32.2 (6.8) | 31.1 (6.8) | 31.1 (7.2) | 29.8 (5.4) | 31.6 (6.2) | 30.2 (6.5) | 29.8 (5.6) | 29.7 (6.6) | 30.3 (5.9) | 30.6 (6.9) | 30.7 (6.5) | 31.6 (7.4) | 29.6 (6.8) |
| missing | 0 | 0 | 0 | 0 | 0 | 0 | 0 | 0 | 1 | 0 | 0 | 0 | 0 |
| civil status |  |  |  |  |  |  |  |  |  |  |  |  |  |
| married | 41 (85%) | 25 (86%) | 48 (76%) | 58 (87%) | 56 (81%) | 68 (87%) | 68 (83%) | 45 (76%) | 55 (81%) | 85 (78%) | 90 (78%) | 54 (81%) | 53 (82%) |
| single or missing | 7 (15%) | 4 (14%) | 15 (24%) | 9 (13%) | 13 (19%) | 10 (13%) | 14 (17%) | 14 (24%) | 13 (19%) | 24 (22%) | 26 (22%) | 13 (19%) | 12 (18%) |
| living in Lausanne | 30 (63%) | 17 (59%) | 37 (59%) | 58 (87%) | 44 (64%) | 51 (65%) | 49 (60%) | 37 (63%) | 39 (57%) | 62 (57%) | 63 (54%) | 32 (48%) | 39 (60%) |
| gravidity |  |  |  |  |  |  |  |  |  |  |  |  |  |
| 1 | 6 (13%) | 2 (7%) | 8 (13%) | 11 (16%) | 10 (14%) | 8 (10%) | 12 (15%) | 10 (17%) | 10 (15%) | 19 (17%) | 21 (18%) | 13 (19%) | 10 (15%) |
| 2 | 2 (4%) | 5 (17%) | 8 (13%) | 9 (13%) | 11 (16%) | 11 (14%) | 11 (13%) | 9 (15%) | 10 (15%) | 23 (21%) | 27 (23%) | 16 (24%) | 9 (14%) |
| >2 | 39 (83%) | 22 (76%) | 46 (74%) | 47 (70%) | 48 (70%) | 58 (75%) | 59 (72%) | 40 (68%) | 48 (71%) | 67 (61%) | 68 (59%) | 38 (57%) | 46 (71%) |
| missing | 1 | 0 | 1 | 0 | 0 | 1 | 0 | 0 | 0 | 0 | 0 | 0 | 0 |
| HISCO class |  |  |  |  |  |  |  |  |  |  |  |  |  |
| 1 | 9 (19%) | 6 (21%) | 9 (14%) | 6 (9%) | 8 (12%) | 9 (12%) | 8 (10%) | 8 (14%) | 8 (12%) | 9 (8%) | 8 (7%) | 5 (7%) | 5 (8%) |
| 2 | 34 (71%) | 22 (76%) | 50 (79%) | 59 (88%) | 55 (80%) | 61 (78%) | 65 (79%) | 46 (78%) | 52 (76%) | 87 (80%) | 96 (83%) | 53 (79%) | 55 (85%) |
| 3 | 0 (0%) | 1 (3%) | 1 (2%) | 0 (0%) | 2 (3%) | 1 (1%) | 5 (6%) | 2 (3%) | 5 (7%) | 3 (3%) | 3 (3%) | 2 (3%) | 1 (2%) |
| missing | 5 (10%) | 0 (0%) | 3 (5%) | 2 (3%) | 4 (6%) | 7 (9%) | 4 (5%) | 3 (5%) | 3 (4%) | 10 (9%) | 9 (8%) | 7 (10%) | 4 (6%) |
| gestational age (weeks) | 13.4 (4.8) | 13.8 (5.5) | 13.8 (6.4) | 12.9 (5.5) | 13.7 (6.7) | 15.0 (6.3) | 15.0 (6.2) | 12.9 (5.7) | 13.0 (4.9) | 12.5 (6.7) | 13.3 (6.0) | 14.4 (6.4) | 14.6 (7.9) |
| missing | 27 | 10 | 17 | 20 | 15 | 23 | 13 | 7 | 11 | 21 | 37 | 27 | 16 |
| syphilis | 0 (0%) | 1 (3%) | 2 (3%) | 1 (1%) | 1 (1%) | 0 (0%) | 0 (0%) | 1 (2%) | 1 (1%) | 4 (4%) | 1 (1%) | 4 (6%) | 2 (3%) |
| flu during pregnancy | 0 (0%) | 0 (0%) | 0 (0%) | 0 (0%) | 0 (0%) | 0 (0%) | 0 (0%) | 0 (0%) | 0 (0%) | **14 (13%)** | **45 (39%)** | **39 (58%)** | 0 (0%) |

**Table S21: maternal characteristics and gestational age for miscarriages compared to births between 1918 and 1920.** p: p-value. *^1^*Mean (SD); n (%). *^2^*Wilcoxon rank sum test; Pearson's Chi-squared test

| **Variable** | **deliveries**, *n* = 2,571*^1^* | **miscarriages**, *n* = 292*^1^* | **p***^2^* |
| --- | --- | --- | --- |
| maternal age (years) | 28.5 (6.3) | 30.9 (6.9) | **<0.001** |
| missing | 6 | 0 |  |
| civil status |  |  | **<0.001** |
| married | 2,238 (87%) | 229 (78%) |  |
| single or missing | 333 (13%) | 63 (22%) |  |
| living in Lausanne | 975 (59%) | 157 (54%) | 0.057 |
| gravidity |  |  | <0.001 |
| 1 | 1,021 (40%) | 53 (18%) |  |
| 2 | 557 (22%) | 66 (23%) |  |
| >2 | 993 (39%) | 173 (59%) |  |
| missing |  |  |  |
| gestational age (weeks) | 39.9 (2.9) | 13.2 (6.4) | <0.001 |
| missing | 140 | 85 |  |
| syphilis | 27 (1%) | 9 (3%) | 0.008 |
| flu during pregnancy | 299 (12%) | 98 (34%) | <0.001 |
| HISCO class |  |  | 0.11 |
| 2 | 2,210 (86%) | 236 (81%) |  |
| 1 | 157 (6%) | 22 (8%) |  |
| 3 | 50 (2%) | 8 (3%) |  |
| missing | 154 (6%) | 26 (9%) |  |
